# Etiology and incidence of diarrhea requiring hospitalization in children under 5 years of age in 31 low- and middle-income countries: findings from the Global Pediatric Diarrhea Surveillance network, 2017–2022

**DOI:** 10.64898/2026.07.26.26358975

**Authors:** Heidi M. Soeters, Sébastien Antoni, Shilpa S. Iyer, Goitom Weldegebriel, Joseph Biey, Jason M. Mwenda, Gloria Rey-Benito, Claudia Ortiz, Roberta Pastore, Dovile Videbaek, Simarjit Singh, Emmanuel Njambe, Lucky Sangal, Deepak Dhongde, Varja Grabovac, Josephine Logronio, Kamal Fahmy, Amany Ghoniem, George Armah, Francis E. Dennis, Mapaseka L. Seheri, Nonkululeko Magagula, Kebareng Rakau-Nondela, Tulio M. Fumian, Irene T.A. Maciel, Elena Samoilovich, Galina Semeiko, Tintu Varghese, Sarah Thomas, Julie Bines, Dandi Li, Furqan Kabir, Jie Liu, Eric R. Houpt, Rashi Gautam, Sara A. Mirza, Jan Vinjé, Mick N. Mulders, Jacqueline E. Tate, Umesh D. Parashar, James A. Platts-Mills, Global Pediatric Diarrhea Surveillance network

**Author notes:** Membership in the Global Pediatric Diarrhea Surveillance network is provided in the Appendix of https://www.medrxiv.org/content/10.64898/2026.05.21.26352576v1.

## Abstract

**Background:** Diarrhea remains a leading cause of child morbidity and mortality. Improved and ongoing estimates of the etiology of hospitalized pediatric diarrhea in low- and middle-income countries (LMICs) are needed to help prioritize and evaluate the use of existing and upcoming vaccines and interventions.

**Methods:** The Global Pediatric Diarrhea Surveillance (GPDS) network is a World Health Organization (WHO)-coordinated public health surveillance network investigating the etiology of hospitalized diarrhea among children aged <5 years in LMICs. The GPDS network enrolls children hospitalized with diarrhea at 38 sentinel surveillance sites in 31 LMICs. Randomly selected stool specimens were tested by TaqMan Array Card quantitative reverse-transcription polymerase chain reaction (qPCR) for 16 pathogens associated with diarrhea. We estimated pathogen-specific attributable fractions (AFs) and incidence of diarrheal hospitalizations at the global, regional, and country levels during 3 time periods: 2017-2018, 2019-2020, and 2021-2022, with a focus on the most recent results.

**Results:** During 2017–2022, the GPDS network enrolled 70,750 children aged <5 years hospitalized with diarrhea, of which 16,458 (23.3%) were randomly selected for qPCR testing. The most prevalent pathogen detected, regardless of quantity or modeled etiologic attribution, was rotavirus (weighted prevalence 30.4%), followed by adenovirus 40/41 (19.1%), norovirus (17.9%), *Shigella* (14.1%), and *Campylobacter jejuni/coli* (8.6%). Overall, in 2017-2022, rotavirus was the leading etiology globally (AF 32.5%; 95% Confidence Interval (CI): 27.4, 37.6), followed by *Shigella* (9.8%; 8.4, 11.3), adenovirus 40/41 (8.6%; 6.3, 10.8) and norovirus (6.7%; 5.6, 7.7). Over time, rotavirus consistently declined from an AF of 36.7% (95% CI: 28.7, 46.7) in 2017-2018 to 26.7% (20.7, 34.1) in 2021-2022. Norovirus AF increased slightly from 6.2% (4.7, 7.7) in 2017-2018 to 7.3% (5.0, 9.2) in 2021-2022. Global *Shigella* burden remained stable, and adenovirus 40/41 demonstrated significant volatility, peaking globally in 2019-2020 (11.9%; 5.9, 17.5). In 2021-2022, rotavirus was the leading cause of hospitalized diarrhea in 6 of 9 geographic groupings, norovirus predominated in Central and South America, and *Shigella* was the leading etiology in South Asia. In the subset of countries that had introduced rotavirus vaccine, the leading etiologies in 2021-2022 were rotavirus (18.4%; 15.8, 21.5) and *Shigella* (16.4%; 11.8, 21.1). In 2021-2022, rotavirus had the highest attributable incidence of hospitalized diarrhea in children (2.3 per 1,000 child-years; 1.8, 3.0), followed by *Shigella* (0.9; 0.7, 1.1), norovirus (0.6; 0.4, 0.8) and adenovirus 40/41 (0.6; 0.4, 0.8).

**Conclusions:** Despite the widespread use of rotavirus vaccines, rotavirus remained the leading cause of severe diarrhea among children aged <5 years in LMICs globally. However, the proportion of pediatric diarrhea attributable to rotavirus consistently declined from 2017-2018 to 2021-2022, and there were notable differences in the distribution of diarrheal etiologies between regions and across time periods. *Shigella*, norovirus, and enteric adenoviruses were also associated with a substantial burden of disease. Improving the efficacy and coverage of rotavirus vaccination and prioritizing interventions against other enteric pathogens could further reduce diarrhea morbidity and mortality.

## Introduction

Recent improvements in global life expectancy have resulted from progress in the prevention and treatment of enteric diseases across the lifespan, in particular diarrheal diseases (1,2). However, diarrhea remains a leading cause of death and disease in children aged <5 years, causing an estimated 340,000 to 440,000 childhood deaths in 2021, predominantly in sub-Saharan Africa and South and Southeast Asia (2,3). In particular, 24% to 35% of the deaths in this youngest age group are estimated to be caused by rotavirus (2,3).

As of April 2026, 140 countries have introduced rotavirus vaccines into their national immunization programs, with average global coverage of the last dose at 59% in 2024 (4,5). Rotavirus vaccines have decreased severe rotavirus diarrhea in children by approximately 40% in low- and middle-income countries (LMICs) (6,7), decreased global rotavirus hospitalizations by 59% (8), and have prevented an estimated 139,000 childhood deaths globally (9). In this context of increasing rotavirus vaccine use and impact, ongoing surveillance of the burden and etiology of rotavirus and other infectious causes of pediatric diarrhea is critical to continue monitoring the benefits of rotavirus vaccines, optimize their use, and inform development of vaccines targeting other pediatric diarrheal pathogens.

The World Health Organization (WHO) has coordinated the Global Rotavirus Surveillance Network (GRSN) since 2008, providing a global source of rotavirus surveillance data among hospitalized children aged <5 years to help monitor rotavirus disease burden and circulating strains, and to evaluate rotavirus vaccine impact (6). However, systematically collected and analyzed surveillance data on the other etiologies of diarrhea in hospitalized children has been limited, particularly in LMICs where the burden is highest (2).

In 2017, WHO leveraged the GRSN sentinel-site surveillance and regional reference laboratory (RRL) system to establish Global Pediatric Diarrhea Surveillance (GPDS) (10). GPDS investigates the etiology of diarrhea among hospitalized children aged <5 years by using a standardized protocol for quantitative reverse-transcription PCR (qPCR) testing for a broad panel of diarrheal pathogens using TaqMan Array Cards. GPDS aims to identify pathogens associated with children hospitalized with diarrhea, including monitoring the changing burden of pathogens over time, monitoring circulating strains, and generating country-level data to inform decision-making around public health interventions. GPDS also provides data that are used to improve global, regional, and country diarrheal disease burden estimates, inform new enteric vaccine development, and GPDS could potentially provide a platform for future enteric vaccine evaluation.

The GPDS network includes sentinel surveillance sites in 31 LMICs across all 6 WHO Regions (10). Initial results from cases enrolled during 2017–2018 demonstrated that despite the substantial impact of rotavirus vaccine introduction, rotavirus remained the leading cause of pediatric diarrhea hospitalizations (11). However, the difference in the proportion of attributable disease between rotavirus and other pathogens closed substantially in countries that had introduced rotavirus vaccine, and countries that had introduced rotavirus vaccine had a lower burden of rotavirus than countries that had not yet introduced the vaccine. Notably, *Shigella*, norovirus, and adenovirus 40/41 were other important causes of diarrhea requiring hospitalization, with substantial geographic heterogeneity in the relative burden of specific pathogens. Here, we report updated global, regional, and country-level findings from the GPDS network from 2017 through 2022, with a focus on the most recent results from 2021-2022 and how the findings have changed since 2017-2018 and 2019-2020.

## Methods

Detailed methods for Global Pediatric Diarrhea Surveillance are described separately (10). Briefly, GPDS sentinel surveillance hospitals were selected among GRSN sites that enrolled a minimum of 100 cases per year of hospitalized diarrhea in children aged 0-59 months. GPDS sites prospectively enroll all children admitted for diarrhea, regardless of symptom duration or the presence of blood in the stool. Diarrhea was defined as 3 or more loose stools in a 24-hour period. An acute diarrhea episode was defined as an illness with a duration prior to enrollment of fewer than 14 days (through a threshold of <7 days was used in some participating countries), while longer episodes were considered persistent.

Thirty-eight sentinel surveillance sites in 31 countries across all 6 WHO regions participated in GPDS in 2017 through 2022 (Fig 1). Some countries had only one surveillance site with all patients enrolled from a single sentinel surveillance hospital. Other countries aggregated patients from two or three surveillance hospitals and reported as a single site. Three large countries had multiple GPDS surveillance sites in geographically diverse areas: China (3 sites), India (5 sites) (12), and Pakistan (2 sites).

**Fig 1.**
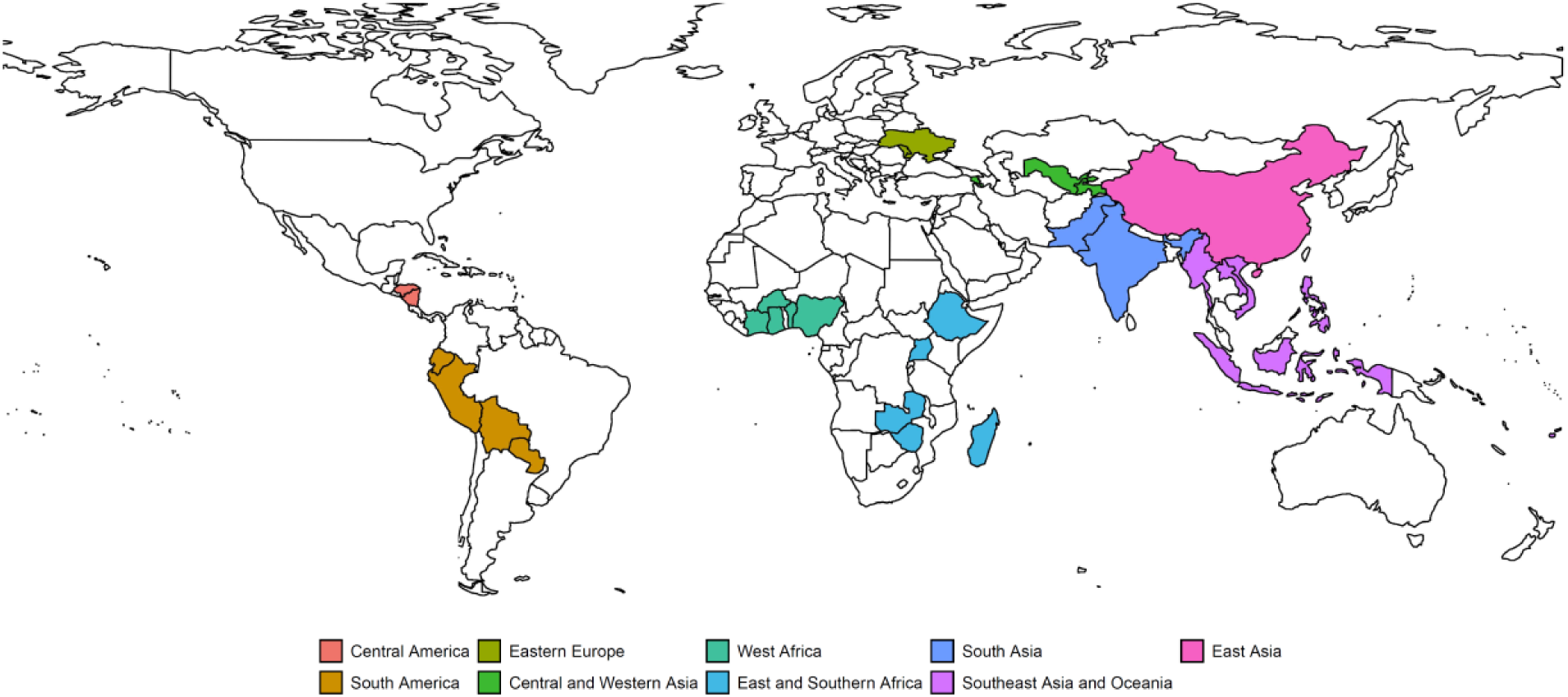
The 38 surveillance sites from 31 countries in the Global Pediatric Diarrhea Surveillance network in 2017-2022. The map indicates the country locations as well as their geographic groupings.

From each surveillance site, 100 cases were randomly selected for qPCR testing and were shipped to a RRL (10). Sample were tested using custom-designed TAC qPCR cards (Thermo Fisher, Waltham, MA, USA) (10), which included qPCR assays for the 16 enteric pathogens that were associated with pediatric diarrhea in two prior multisite studies that incorporated diarrheal cases and non-diarrheal controls: the Global Enteric Multicenter Study (GEMS) and the Etiology, Risk Factors, and Interactions of Enteric Infections and Malnutrition and the Consequences for Child Health and Development (MAL-ED) birth cohort study (13,14). Detections with a cycle threshold (Ct) value less than 35 were considered positive.

For analysis, we applied inverse probability of selection weights such that cases selected for testing were representative of all enrolled cases, including appropriately capturing seasonal variation in diarrhea etiology (10). As asymptomatic identification of enteropathogens is common in LMICs, especially when using molecular testing methods, we attributed diarrhea to specific enteropathogens based on pathogen quantity using models developed from qPCR re-analyses of the GEMS case-control study and the MAL-ED birth cohort study (13–15). Weighted population attributable fractions (AFs) for each pathogen were calculated and then applied to national-level estimates of the number of both diarrhea hospitalizations and children aged <5 years from the Global Burden of Disease (GBD) 2021 study to estimate the pathogen-specific incidence of hospitalized diarrhea (16). Estimates were produced at the country, regional, and global level in two-year analytic groupings (2017-2018, 2019-2020, and 2021-2022) and overall for the entire 2017–2022 period for select estimates. Analytic geographic groupings were created to provide greater distinction and public health inferences for the GPDS estimates than the 6 WHO Regions; see Fig 1 for the geographic grouping assigned to each country and surveillance site. Additionally, a sub-analysis produced estimates limited to sites that had introduced rotavirus vaccine to the immunization program at the country or regional level by the end of first year of each two-year analytic grouping. For all analyses, point estimates and 95% confidence intervals were derived from the median and 2.5th and 97.5th quantiles respectively of the estimate distributions. All analyses were conducted in R version 4.0.2. Analyses were run using GPDS surveillance data available as of 16 January 2026.

### Ethical considerations

Surveillance activities that are part of GPDS are exempt from research ethical review by WHO, since they are considered public health surveillance (17). Authors had no access to information that could identify individual participants during or after data collection.

## Results

From January 2017 to December 2022, 70,750 children aged <5 years hospitalized with diarrhea were enrolled from 38 surveillance sites in 31 countries (Fig 1; Table 1; and (10)). Using GBD diarrheal mortality and population estimates for all LMICs in 2021, these 31 countries represented 75.9% (246,647/324,946) of all diarrheal deaths and 68.0% (329,133,825/484,384,297) of the population of children aged <5 years.

**Table 1.** Demographic and clinical characteristics of children aged <5 years hospitalized with diarrhea enrolled in Global Pediatric Diarrhea Surveillance, 2017-2022.

|  | Total |  | 2017–2018<sup>a</sup> |  | 2019–2020 |  | 2021–2022 |  |
| --- | --- | --- | --- | --- | --- | --- | --- | --- |
| Demographic and clinical characteristics | All enrolled cases<br>N = 70750 | qPCR-tested cases<sup>b</sup><br>N = 16458<br>(23.3) | All enrolled cases<br>N = 31100 | qPCR-tested cases<sup>b</sup><br>N = 5670<br>(18.2) | All enrolled cases<br>N = 19367 | qPCR-tested cases<sup>b</sup><br>N = 5509<br>(28.4) | All enrolled cases<br>N = 20283 | qPCR-tested cases<sup>b</sup><br>N = 5279<br>(26.0) |
| Male sex | 41005 (58.0) | 9578 (58.3) | 18073 (58.2) | 3341 (58.7) | 11194 (57.9) | 3218 (59.1) | 11738 (57.9) | 3019 (56.8) |
| Age |  |  |  |  |  |  |  |  |
| 0-11 months | 31848 (45.0) | 7237 (45.7) | 13706 (44.1) | 2512 (44.8) | 9015 (46.5) | 2478 (47.3) | 9127 (45.0) | 2247 (45.5) |
| 12-23 months | 22791 (32.2) | 5300 (32.1) | 10356 (33.3) | 1848 (33.0) | 6210 (32.1) | 1800 (32.2) | 6225 (30.7) | 1652 (30.7) |
| 24-59 months | 16111 (22.8) | 3921 (22.2) | 7038 (22.6) | 1310 (22.2) | 4142 (21.4) | 1231 (20.5) | 4931 (24.3) | 1380 (23.7) |
| Acute diarrhea (<14 days) | 69758 (98.6) | 16218 (98.7) | 30651 (98.6) | 5587 (98.7) | 19055 (98.4) | 5415 (98.4) | 20052 (98.9) | 5216 (99.0) |
| Diarrheal duration (days) | 3 (2, 4) | 3 (2, 4) | 3 (1, 4) | 3 (1, 4) | 3 (2, 4) | 3 (2, 4) | 3 (2, 4) | 3 (2, 4) |
| Bloody diarrhea | 4097 (6.7) | 1016 (6.5) | 1771 (6.5) | 276 (5.6) | 1316 (7.1) | 402 (7.3) | 1010 (6.4) | 338 (7.1) |
| Vomiting | 46830 (68.4) | 10958 (68.9) | 21130 (70.6) | 3719 (71.6) | 12072 (65.0) | 3665 (66.0) | 13628 (68.1) | 3574 (67.6) |
| Dehydration | 40927 (75.3) | 8145 (76.4) | 19038 (72.9) | 2779 (70.7) | 10482 (74.0) | 2766 (80.5) | 11407 (81.0) | 2600 (82.1) |
| Rehydration therapy given | 39998 (74.7) | 7914 (76.1) | 18593 (72.4) | 2713 (70.4) | 10184 (73.5) | 2650 (80.1) | 11221 (80.3) | 2551 (81.8) |
| In-hospital deaths | 319 (0.5) | 90 (0.4) | 157 (0.5) | 39 (0.4) | 73 (0.4) | 24 (0.4) | 89 (0.5) | 27 (0.5) |
| World Health Organization Region |  |  |  |  |  |  |  |  |
| African | 20903 (29.5) | 5325 (29.3) | 9327 (30.0) | 1827 (29.6) | 5425 (28.0) | 1768 (27.7) | 6151 (30.3) | 1730 (30.5) |
| Americas | 12392 (17.5) | 3131 (17.6) | 5428 (17.5) | 1010 (17.5) | 3594 (18.6) | 1112 (18.6) | 3370 (16.6) | 1009 (16.6) |
| Eastern Mediterranean | 4340 (6.1) | 699 (6.2) | 0 (0) | 0 (0) | 731 (3.8) | 298 (3.8) | 3609 (17.8) | 401 (17.8) |
| European | 15685 (22.2) | 2231 (22.3) | 9164 (29.5) | 1000 (29.7) | 3728 (19.2) | 659 (19.5) | 2793 (13.8) | 572 (13.8) |
| South-East Asian | 6137 (8.7) | 2171 (8.7) | 3241 (10.4) | 881 (10.4) | 2147 (11.1) | 735 (11.1) | 749 (3.7) | 555 (3.7) |
| Western Pacific | 11293 (16.0) | 2901 (16.0) | 3940 (12.7) | 952 (12.8) | 3742 (19.3) | 937 (19.3) | 3611 (17.8) | 1012 (17.6) |

Dichotomous estimates are shown as N (%) and continuous estimates are shown as median (interquartile range).
<sup>a</sup>Due to ongoing updates to surveillance data and adjustments to the weighting model, data displayed for 2017-2018 do not exactly match those previously published by Cohen & Platts-Mills et al (11).
<sup>b</sup>Counts are unweighted; percentages are weighted and calculated from the subset of participants with non-missing data for each characteristic.

Of the 70,750 enrolled cases, 68,302 (96.5%) had a stool sample collected and were eligible for qPCR testing; of these, 16,458 (24.1%) were selected and tested: 5,670 from 2017-2018, 5,509 from 2019-2020, and 5,279 from 2021-2022 (Table 1). Most enrolled children were aged <2 years (54,639, 77.8%), and 41,005 (58.3%) were male. Most cases were accompanied by vomiting and dehydration, and almost all patients received some form of rehydration therapy in the hospital. Of 61,480 (86.9%) cases with information recorded on diarrhea duration and presence of blood in stool, 56,570 (92.0%) presented with acute watery diarrhea, 3,992 (6.5%) with acute bloody diarrhea, and 918 (1.5%) with watery or bloody persistent diarrhea. Of 66,881 cases with a discharge outcome available, 319 (0.5%) died during the hospitalization. The demographic and clinical characteristics of the qPCR-tested cases were similar to those characteristics for all cases enrolled in GPDS after application of inverse probability of selection weights.

Overall, a mean of 1.3 (standard deviation [SD] 1.2) pathogens was detected per sample. By region, the mean number of pathogens detected per sample ranged from a low of 0.7 (SD 0.7) in East Asia to a high of 1.8 (SD 1.4) in South Asia. Among all samples tested by qPCR, the most prevalent pathogen detected, regardless of quantity or modeled etiologic attribution, was rotavirus (weighted prevalence 30.4%), followed by adenovirus 40/41 (19.1%), norovirus (17.9%), *Shigella* (14.1%), and *Campylobacter jejuni/coli* (8.6%), with substantial variation by site (S1 Fig). Overall, 63.9% of hospitalized diarrhea was attributable to one of the 16 pathogens included in this analysis, ranging from 34.3% in East Asia to 68.6% in East Africa.

Across the entire period of 2017 to 2022, rotavirus was the leading etiology globally (AF 32.5%; 95% Confidence Interval [CI]: 27.4, 37.6), followed by *Shigella* (9.8%; 8.4, 11.3), adenovirus 40/41 (8.6%; 6.3, 10.8) and norovirus (6.7%; 5.6, 7.7). Rotavirus exhibited a large and consistent decline throughout the period of analysis, with the overall AF falling from 36.7% (95% CI: 28.7, 46.7) in 2017-2018 to 26.7% (20.7, 34.1) in 2021-2022, including a net decrease in all 9 geographic regions (Figs 2 & 3; S1 Table). In contrast, the global burden of *Shigella* remained stable, starting at 10.6% (8.0, 13.6) in 2017-2018 and ending at 10.1% (7.7, 12.6) in 2021-2022. Adenovirus 40/41 demonstrated the most significant volatility, with the overall AF increasing substantially from 6.9% (4.7, 9.0) in 2017-2018 to 11.9% (5.9, 17.5) in 2019-2020, an increase observed in 7 out of 9 regions. However, in 2021-2022 the overall adenovirus 40/41 AF returned to 7.0% (4.1, 9.7), with stepwise declines observed in 8 out of 9 regions; this included a striking reduction in South Asia, where the AF fell from a high of 17.1% (7.5, 27.8) in 2017-2018 to 4.8% (2.6, 7.4) in 2021-2022. Finally, norovirus showed a small but consistent upward trend, increasing from 6.2% (4.7, 7.7) in 2017-2018 to 7.3% (5.0, 9.2) in 2021-2022, with net increases recorded in 5 out of 9 regions.

**Fig 2.**
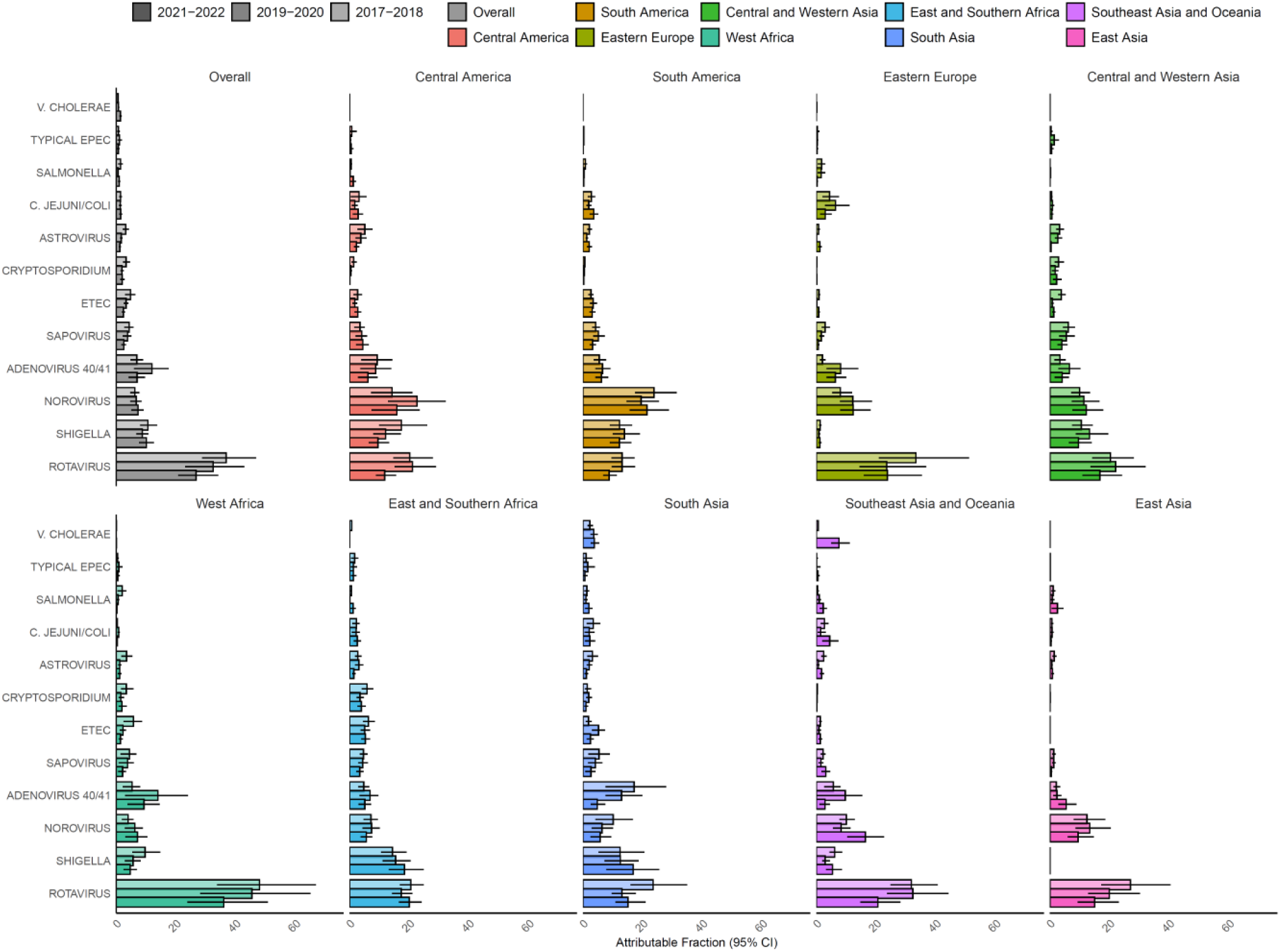
Pathogen-specific attributable fractions of hospitalized diarrhea in children aged <5 years in 2017-2022 in the Global Pediatric Diarrhea Surveillance network both overall and by geographic region. Within each grouping and pathogen, the attributable fractions were weighted by the site-level attributable incidence of hospitalized diarrhea. Bar shadings indicate the two-year periods of surveillance. Attributable fractions are expressed as a percent. EPEC=enteropathogenic *E. coli*; ETEC=enterotoxigenic *E. coli*. Data behind this figure are displayed in S1 Table.

**Fig 3.**
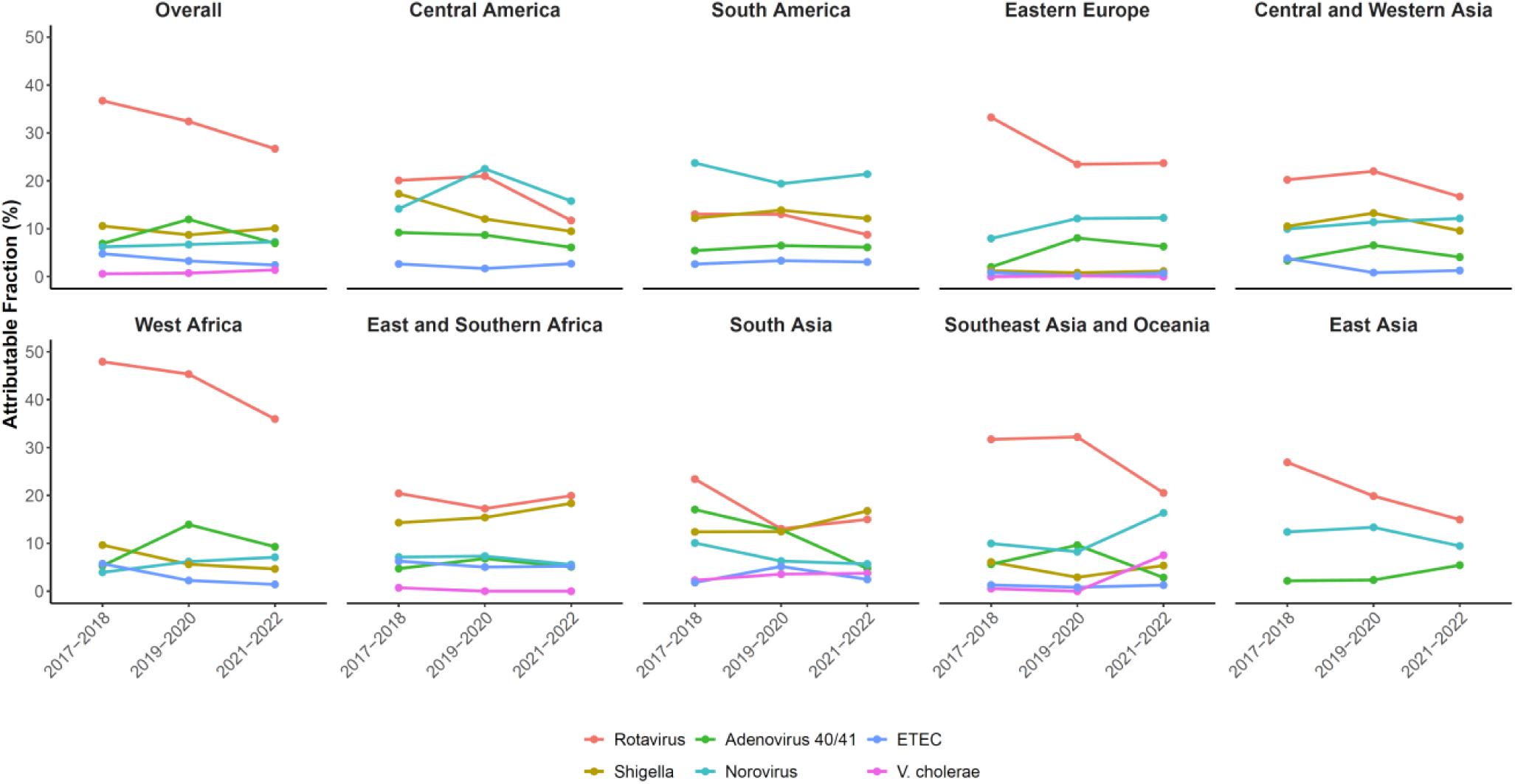
Trends in pathogen-specific attributable fractions from 2017 to 2022 in the Global Pediatric Surveillance Network, both overall and by geographic region. Attributable fractions are shown for each two-year time period: 2017-2018, 2019-2020, and 2021-2022. Pathogens included are those that ranked in the top two for at least one region during at least one time period; within each regional panel, only pathogens with an attributable fraction >0.1% for at least one two-year period are displayed. ETEC=enterotoxigenic *E. coli*. Data behind this figure are displayed in S1 Table.

Looking at the most recent 2021-2022 data by region, rotavirus was the leading cause of hospitalized diarrhea in 6 of 9 regions (AFs ranged from 15.0% in East Asia to 36.0% in West Africa) (Fig 2; S1 Table). Norovirus was the leading etiology in both Central America (AF 15.8%; 7.4, 23.4) and South America (AF 21.4%; 15.6, 28.7). *Shigella* was the leading etiology in South Asia (AF 16.8%; 7.8, 25.5). In the subset of countries that had introduced rotavirus vaccine prior to 2021, the top etiologies in 2021-2022 were rotavirus (18.4%; 15.8, 21.5), *Shigella* (16.4%; 11.8, 21.1), norovirus (6.0%; 4.4, 7.9) and adenovirus 40/41 (5.7%; 4.2, 7.3) (Fig 4; S2 Table).

**Fig 4.**
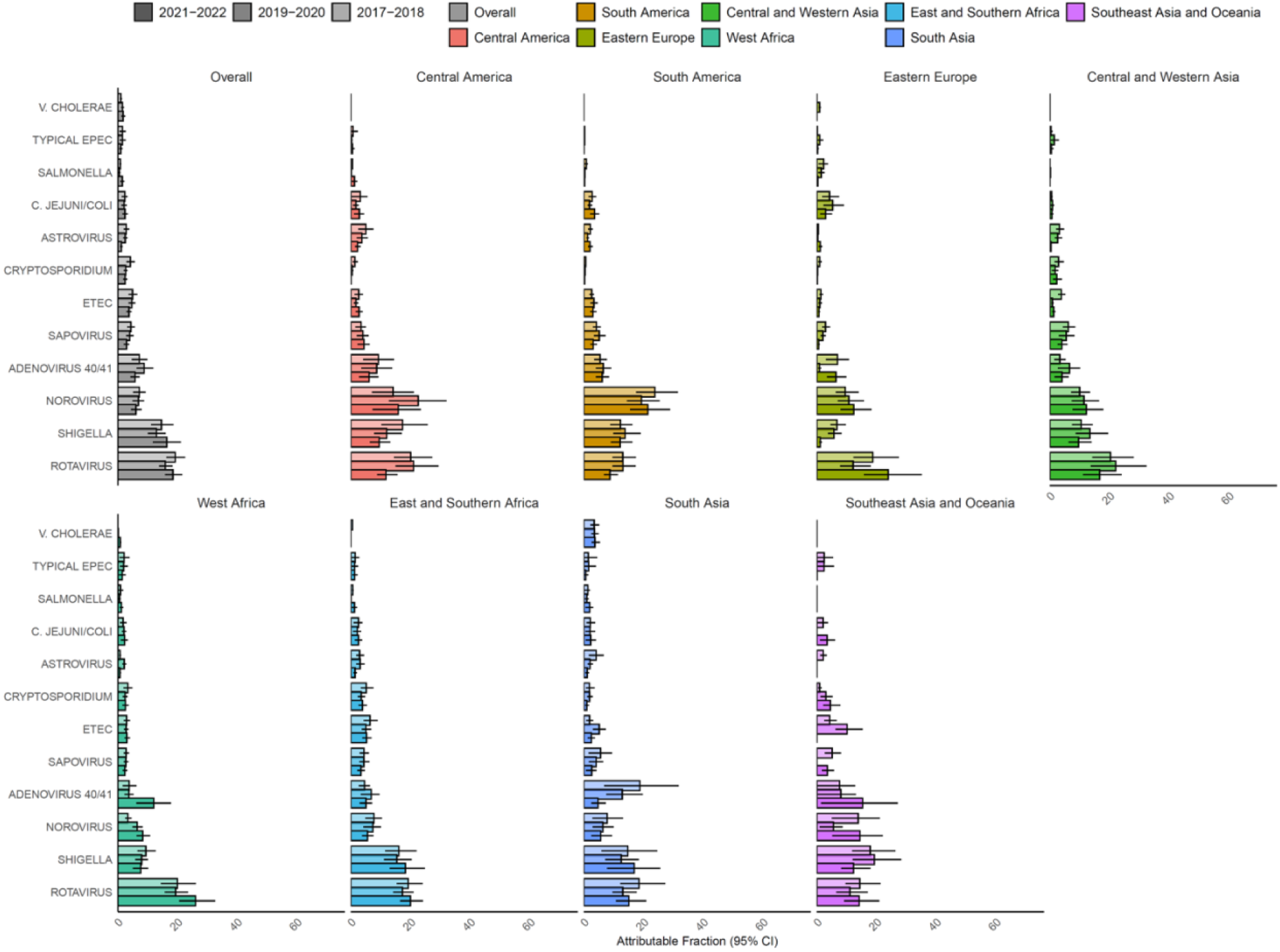
Pathogen-specific attributable fractions of hospitalized diarrhea in children aged <5 years in 2017-2022 in the Global Pediatric Diarrhea Surveillance network both overall and by geographic region, limited to sites that had introduced rotavirus vaccine. Within each grouping and pathogen, the attributable fractions were weighted by the site-level attributable incidence of hospitalized diarrhea. Bar shadings indicate the two-year periods of surveillance. Attributable fractions are expressed as a percent. EPEC=enteropathogenic *E. coli*; ETEC=enterotoxigenic *E. coli.* Data behind this figure are displayed in S2 Table.

A wide variety in diarrhea etiology was observed by site (Fig 5; Tables S3, S4, and S5). For example, some of the highest rotavirus AFs in 2021-2022 were observed in surveillance sites in Zambia, Nigeria, Ghana, and Madagascar. Norovirus was the leading etiology at many sites in Central and South America. The highest AFs for *Shigella* were observed in Madagascar and Ethiopia, and *Shigella* was detected at every surveillance site in 2021-2022 except for the 3 sites in China (S1 Fig). *V. cholerae* was strikingly high in both sites in Pakistan (AFs 17.9% and 9.8%) and in the Philippines (AF 12.6%; 95% CI: 11.3, 13.0); *V. cholerae* was a less common cause of hospitalized diarrhea in Benin and two of the sites in India and was not detected in the other 26 sites in 2021-2022. The highest site-level AFs for the remaining top pathogens were: Adenovirus 40/41 (17.0%) in Ghana, enterotoxigenic *E. coli* (12.4%) in Madagascar, sapovirus (8.9%) in Tirupati, India, *Cryptosporidium* (7.7%) in Uganda, *Campylobacter jejuni/coli* (7.4%) in Mauritius, salmonella (6.3%) in Fuzhou, China, astrovirus (4.9%) in Armenia, and typical enteropathogenic *E. coli* (2.4%) in Uganda and in Karachi, Pakistan.

**Fig 5.**
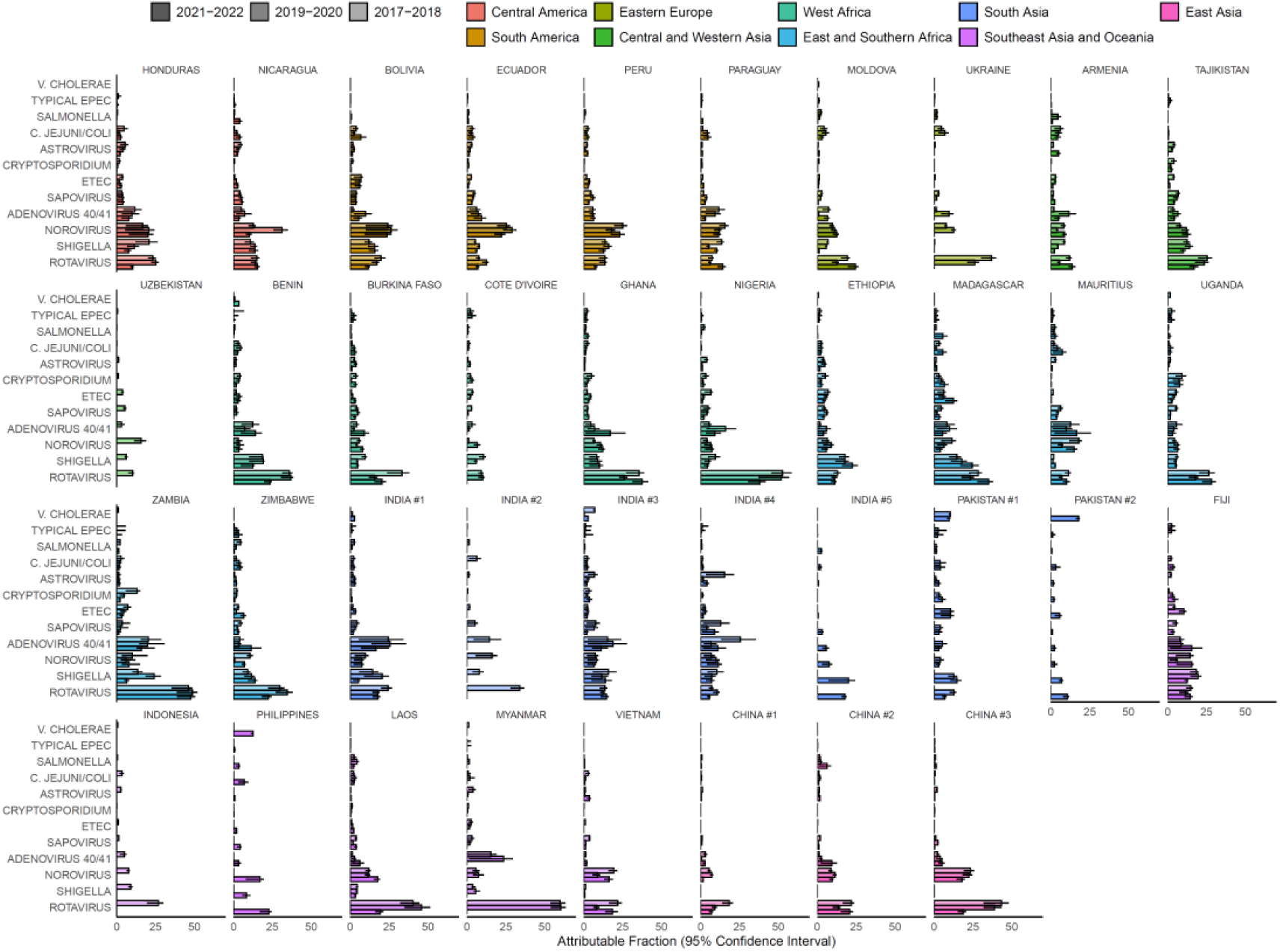
Pathogen-specific attributable fractions of hospitalized diarrhea in children aged <5 years in 2017-2022 for the 38 surveillance sites from 31 countries in the Global Pediatric Diarrhea Surveillance network. The bar plots are colored according to the geographic groupings. Bar shadings indicate the two-year periods of surveillance. Attributable fractions are expressed as a percent. EPEC=enteropathogenic *E. coli*; ETEC=enterotoxigenic *E. coli*. Data behind this figure are displayed in Tables S3, S4, and S5.

Incorporating GBD 2021 diarrhea incidence estimates, the overall incidence of all-cause diarrhea hospitalizations in 2021-2022 was 8.8 per 1,000 child-years (95% CI: 7.4, 10.5) and ranged from 0.5 (0.3, 0.8) in East Asia to 29.1 (21.0, 39.6) in West Africa (S6 Table). Rotavirus had the highest attributable incidence of hospitalized diarrhea in children (2.3; 1.8, 3.0), followed by *Shigella* (0.9; 0.7, 1.1), norovirus (0.6; 0.4, 0.8), and adenovirus 40/41 (0.6; 0.4, 0.8) (Fig 6; S6 Table). Overall, rotavirus had the largest decline in attributable incidence from 4.4 (3.5, 5.6) in 2017-2018 to 2.3 (1.8, 3.0) in 2021-2022.

**Fig 6.**
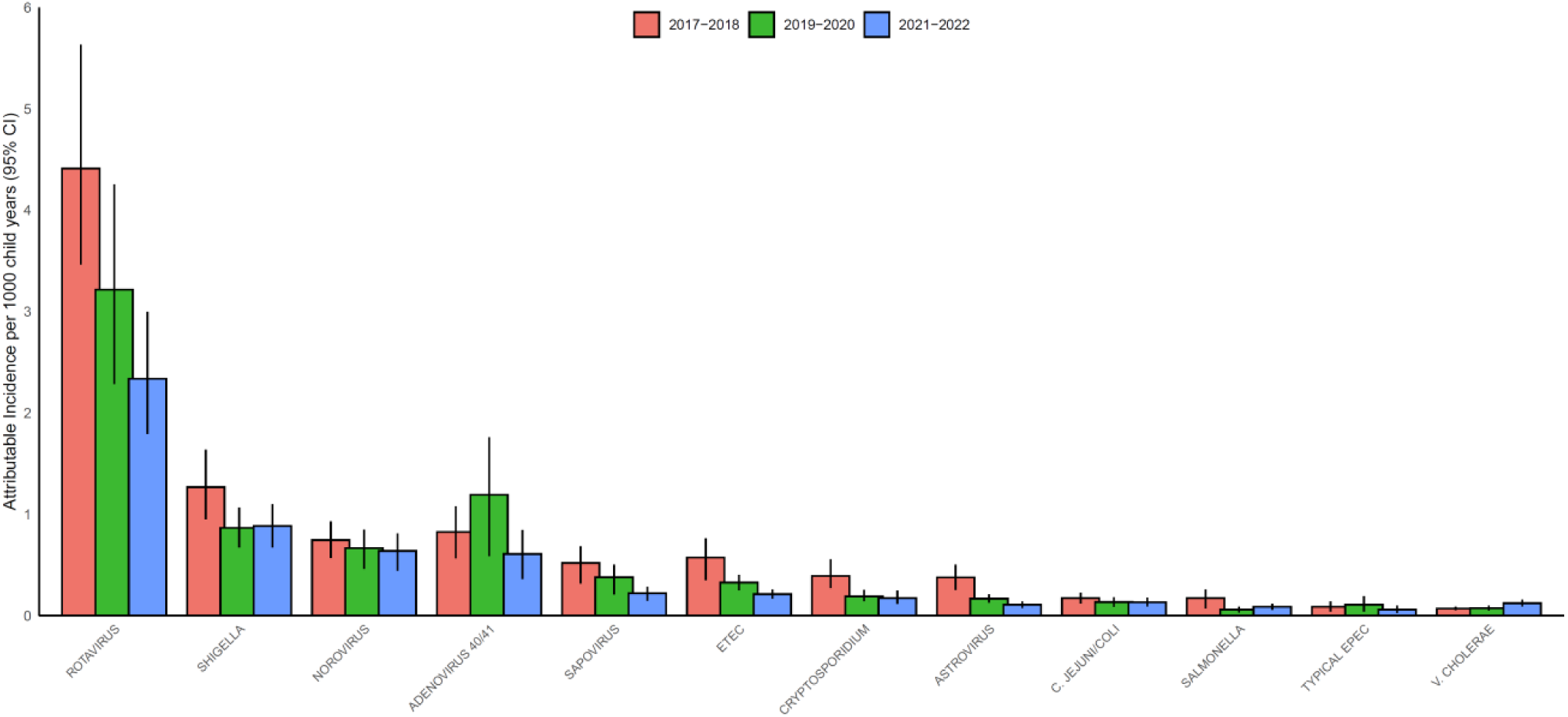
Pathogen-specific attributable incidence of diarrhea hospitalizations in children aged <5 years in 2017-2022 in the Global Pediatric Diarrhea Surveillance network. ETEC=enterotoxigenic *E. coli.* Data behind this figure are displayed in S6 Table.

## Discussion

Despite the widespread use of rotavirus vaccines, rotavirus remained the leading cause of hospitalized diarrhea among children aged <5 years in LMICs globally, including in the subset of countries that had introduced rotavirus vaccine. However, the proportion of pediatric diarrhea attributable to rotavirus consistently declined from 2017-2018 to 2021-2022, and there were notable differences in the distribution of diarrheal etiologies between regions and across time periods. *Shigella*, norovirus, and enteric adenoviruses were also associated with a substantial burden of disease. Improving the efficacy and coverage of rotavirus vaccination and prioritizing interventions against other enteric pathogens could further reduce diarrhea morbidity and mortality.

From 2017-2022, hospitalizations due to rotavirus decreased in both attributable fraction and incidence in all 9 geographic regions. Ongoing monitoring of rotavirus via GPDS, GRSN (6), and other surveillance efforts will continue to measure changes in incidence, especially in countries that introduced the vaccine in recent years, such as Nigeria (4). Among the subset of countries with rotavirus vaccine, 5 of 9 regions experienced an increase in rotavirus AF in 2021-2022 as compared to 2019-2020. This may have been due to a variety of factors, including rotavirus vaccine product disruptions, COVID-19 pandemic impacts on disease transmission and surveillance, and some changes in GPDS site participation over time.

GPDS data have quantified a consistently high burden of norovirus in the Americas, the leading cause of diarrhea hospitalizations in a region that introduced rotavirus vaccines over a decade ago with good uptake; this finding is consistent with previous reports (18–20). A high burden of *Shigella* was detected across a number of GPDS surveillance sites, suggesting that shigellosis is endemic among young children in many LMICs and that a *Shigella* vaccine may have a large and broad impact (21,22). GPDS estimates of adenovirus 40/41 were relatively dynamic across both space and time, indicating variable endemicity and providing valuable data regarding adenovirus 40/41 epidemiology in LMICs (23). Cholera was a major cause of hospitalized diarrhea in a small number of sites, although we acknowledge the limitations of routine sentinel surveillance in detecting outbreaks. These data support the importance of targeted cholera vaccination efforts (24) and the ongoing clinical development of *Shigella* and norovirus vaccines (22,25), as well the need for foundational research to understand adenovirus immunity and potential vaccine targets.

The global COVID-19 pandemic occurred during the time period of this analysis, with substantial and documented impacts on surveillance, public health infrastructure, circulating pathogens, and health care seeking behavior (26,27). Although many efforts were made to sustain GPDS surveillance during the pandemic, and GPDS has a relatively forgiving study design of banking and retrospective testing of stool samples (10), a number of sites experienced surveillance interruptions. To help even out some of this variation, data was analyzed in two-year groupings (2017-2018, 2019-2020, and 2021-2022), with the years most heavily impacted by the COVID-19 pandemic (2020 and 2021) being grouped with 2019 and 2022, respectively. With this caveat, we did not observe a clear shift in the pathogen distribution in the early years of the COVID-19 pandemic, contrary to reports from mainly high-income countries that described a decrease in gastrointestinal infections, particularly viral infections (28–30). The lack of a clear COVID-19 impact in GPDS data may be partially attributed to different settings (LMICs vs high-income countries), differential implementation and effectiveness of non-pharmaceutical public health interventions across sites (31), exclusive focus on severe hospitalized diarrhea, and producing biannual estimates rather than looking at finer temporal gradations.

Strengths of the GPDS design and network have been detailed elsewhere (10). Firstly, GPDS leverages existing surveillance infrastructure to capture information on 16 enteric pathogens, an approach which supports WHO’s global strategy for comprehensive vaccine-preventable disease surveillance (32). Moving beyond single-pathogen siloed surveillance systems promotes efficiency, innovation, and adaptability and produces valuable data for a wider variety of stakeholders. Additionally, this analysis highlighted substantial pathogen diversity across time and between surveillance sites. Continuous surveillance is critical for capturing temporal trends; and in large countries, GPDS includes multiple surveillance sites which have demonstrated substantial within country variability – documenting this heterogeneity is key to informing tailored public health interventions.

Some limitations specific to this analysis include that only 34.3% of hospitalized pediatric diarrhea in East Asia (specifically China) was attributable to one of the pathogens included in GPDS testing, roughly half of the total attribution in some regions; pathogen prevalence at any quantity (without application of the attribution modeling) was also lower in this region. As the GPDS qPCR panel was developed with a global perspective, it may be missing predominant pathogens or strains in certain regions. Additional studies on pediatric diarrheal etiology in these settings could help inform the development of more tailored multiplex diagnostics, especially affordable diagnostics needed for country-led sustainable surveillance.

Variations in the proportion of cases with attributable pathogens identified via GPDS testing could also be partially due to surveillance artifacts such as stool sample quality. Additionally, this analysis does not account for mixed infections, as multiple pathogens are often detected in the same stool sample; mixed infections will be explored in-depth in future analyses. Lastly, because the majority of the GPDS sites are in urban settings, our estimates may be biased towards pathogens that infect via person-to-person transmission, rather than through environmental pathways (33).

By incorporating quantitative molecular diagnostics and pathogen attribution modelling into a global network with standardized surveillance, sample collection and testing, and analytic protocols, GPDS provides direct estimates of pathogens causing hospitalized diarrhea in children aged <5 years in a large, globally representative set of LMICs in 2017–2022. This directly addresses the critical knowledge gap in the etiology of many children with severe diarrheal disease. This network will continue to produce diarrheal disease burden estimates, monitor the changing burden and strains of these pathogens over time, and generate data to inform public health decision-making. Using these data to improve the efficacy and coverage of rotavirus vaccination, while also prioritizing interventions against other enteric pathogens could further reduce diarrhea morbidity and mortality among children worldwide.

## Data Availability

Aggregate GPDS data is available online at: https://immunizationdata.who.int/global?topic=Rotavirus-and-pediatric-diarrhea-surveillance-data. De-identified participant-level data used in these analyses will be made available by WHO upon request to qualified researchers, after approval of a proposal submitted to and signing of a WHO data sharing agreement.

## Acknowledgements

We would like to thank the sentinel surveillance hospitals, national laboratories, and staff and the country Ministries of Health for supporting and maintaining surveillance at the country level, as well as the WHO Country Offices. The findings and conclusions of this report are those of the authors and do not necessarily represent the official position of the U.S. Centers for Disease Control and Prevention or the World Health Organization.

## Supplemental Material

**S1 Table.** Pathogen-specific attributable fractions with 95% confidence intervals of diarrhea hospitalizations in children aged <5 years both overall and by geographic grouping in countries participating in Global Pediatric Diarrhea Surveillance, by two-year time periods during 2017-2022 (Underlying data for Figs 2 and 3).

**S2 Table.** Pathogen-specific attributable fractions with 95% confidence intervals of diarrhea hospitalizations in children aged <5 years both overall and by geographic grouping in countries participating in Global Pediatric Diarrhea Surveillance, limited to sites that had introduced rotavirus vaccine, by two-year time periods during 2017-2022 (Underlying data for Fig 4).

**S3 Table.** Pathogen-specific attributable fractions with 95% confidence intervals of diarrhea hospitalizations in children aged <5 years by Global Pediatric Diarrhea Surveillance site, 2021-2022 (Underlying data for Fig 5).

**S4 Table.** Pathogen-specific attributable fractions with 95% confidence intervals of diarrhea hospitalizations in children aged <5 years by Global Pediatric Diarrhea Surveillance site, 2019-2020 (Underlying data for Fig 5).

**S5 Table.** Pathogen-specific attributable fractions with 95% confidence intervals of diarrhea hospitalizations in children aged <5 years by Global Pediatric Diarrhea Surveillance site, 2017-2018 (Underlying data for Fig 5).

**S6 Table.** All-cause and pathogen-specific attributable incidence per 1000 child-years with 95% confidence intervals of diarrhea hospitalizations in children aged <5 years both overall and by geographic grouping in countries participating in Global Pediatric Diarrhea Surveillance, by two-year time periods during 2017-2022 (Underlying data for Fig 6).

**S1 Fig.** Prevalence of pathogens including in the attribution modelling at cycle threshold cutoffs of 35 and 30 in children <5 years hospitalized with diarrhea and enrolled in Global Pediatric Diarrhea Surveillance, by site and two-year time period from 2017-2022.

