## Supplementary material for "Etiology and incidence of diarrhea requiring hospitalization in children under 5 years of age in 31 low- and middle-income countries: findings from the Global Pediatric Diarrhea Surveillance network, 2017–2022": S1 Appendix

### Table of Contents

S1 Table. Pathogen-specific attributable fractions with 95% confidence intervals of diarrhea hospitalizations in children aged <5 years both overall and by geographic grouping in countries participating in Global Pediatric Diarrhea Surveillance, by two-year time periods during 2017-2022 (Underlying data for Figures 2 and 3). ............... 2

S2 Table. Pathogen-specific attributable fractions with 95% confidence intervals of diarrhea hospitalizations in children aged <5 years both overall and by geographic grouping in countries participating in Global Pediatric Diarrhea Surveillance, limited to sites that had introduced rotavirus vaccine, by two-year time periods during 2017-2022 (Underlying data for Figure 4). ............... 3

S3 Table. Pathogen-specific attributable fractions with 95% confidence intervals of diarrhea hospitalizations in children aged <5 years by Global Pediatric Diarrhea Surveillance site, 2021-2022 (Underlying data for Figure 5). ... 4

S4 Table. Pathogen-specific attributable fractions with 95% confidence intervals of diarrhea hospitalizations in children aged <5 years by Global Pediatric Diarrhea Surveillance site, 2019-2020 (Underlying data for Figure 5). ... 5

S5 Table. Pathogen-specific attributable fractions with 95% confidence intervals of diarrhea hospitalizations in children aged <5 years by Global Pediatric Diarrhea Surveillance site, 2017-2018 (Underlying data for Figure 5). ... 6

S6 Table. All-cause and pathogen-specific attributable incidence per 1000 child-years with 95% confidence intervals of diarrhea hospitalizations in children aged <5 years both overall and by geographic grouping in countries participating in Global Pediatric Diarrhea Surveillance, by two-year time periods during 2017-2022 (Underlying data for Figure 6). ............... 7

S1 Fig. Prevalence of pathogens including in the attribution modelling at cycle threshold cutoffs of 35 and 30 in children <5 years hospitalized with diarrhea and enrolled in Global Pediatric Diarrhea Surveillance, by site and two-year time period from 2017-2022. ............... 8

**S1 Table. Pathogen-specific attributable fractions with 95% confidence intervals of diarrhea hospitalizations in children aged <5 years both overall and by geographic grouping in countries participating in Global Pediatric Diarrhea Surveillance, by two-year time periods during 2017-2022 (Underlying data for Figures 2 and 3).**

| | Rotavirus | *Shigella* | Norovirus | Adenovirus 40/41 | Sapovirus | ETEC | *Cryptosporidium* | Astrovirus | *C. jejuni/coli* | *Salmonella* | tEPEC | *V. cholerae* |
|---|---|---|---|---|---|---|---|---|---|---|---|---|
| **2017-2018** | | | | | | | | | | | | |
| Overall | 36.7 (28.7, 46.7) | 10.6 (8.0, 13.6) | 6.2 (4.7, 7.7) | 6.9 (4.7, 9.0) | 4.3 (2.7, 5.7) | 4.8 (2.9, 6.3) | 3.3 (2.3, 4.6) | 3.1 (2.1, 4.2) | 1.4 (1.0, 1.9) | 1.4 (0.6, 2.2) | 0.7 (0.3, 1.2) | 0.6 (0.5, 0.7) |
| Central America | 20.1 (14.6, 27.8) | 17.3 (9.8, 25.9) | 14.2 (7.2, 21.0) | 9.2 (3.8, 14.3) | 3.4 (1.5, 5.1) | 2.6 (1.2, 4.1) | 1.3 (0.6, 2.3) | 5.1 (2.6, 7.6) | 3.1 (0.3, 5.6) | 0.5 (0.2, 0.8) | 0.7 (0.0, 2.3) | 0.0 (0.0, 0.0) |
| South America | 13.0 (9.5, 17.2) | 12.2 (8.9, 16.4) | 23.7 (17.4, 31.3) | 5.4 (3.5, 7.7) | 4.2 (3.1, 5.6) | 2.6 (1.8, 3.5) | 0.6 (0.4, 0.8) | 2.1 (1.4, 2.9) | 2.8 (1.6, 4.1) | 0.8 (0.2, 1.4) | 0.2 (0.1, 0.4) | 0.0 (0.0, 0.0) |
| Eastern Europe | 33.2 (20.9, 51.1) | 1.2 (0.9, 1.6) | 8.0 (5.3, 11.9) | 2.0 (1.2, 3.1) | 2.9 (1.8, 4.5) | 0.8 (0.5, 1.2) | 0.1 (0.1, 0.2) | 0.7 (0.4, 1.1) | 4.4 (2.1, 7.5) | 1.7 (0.4, 2.9) | 0.3 (0.0, 0.9) | 0.0 (0.0, 0.0) |
| Central and Western Asia | 20.2 (14.1, 28.0) | 10.5 (7.4, 14.3) | 9.9 (7.1, 13.4) | 3.4 (1.5, 5.3) | 6.2 (4.4, 8.4) | 3.8 (2.7, 5.3) | 2.9 (1.6, 4.7) | 3.2 (2.1, 4.7) | 0.6 (0.3, 0.8) | 0.1 (0.0, 0.2) | 0.4 (0.1, 0.9) | 0.0 (0.0, 0.0) |
| West Africa | 47.9 (33.7, 66.7) | 9.6 (5.4, 14.7) | 3.9 (1.9, 5.7) | 5.3 (2.1, 8.0) | 4.4 (1.5, 6.7) | 5.7 (2.4, 8.6) | 3.4 (1.6, 5.7) | 3.5 (1.7, 5.4) | 0.4 (0.3, 0.5) | 2.0 (0.5, 3.3) | 0.4 (0.1, 0.8) | 0.1 (0.1, 0.1) |
| East and Southern Africa | 20.4 (16.8, 24.8) | 14.3 (10.5, 19.0) | 7.1 (4.7, 9.5) | 4.7 (2.9, 6.5) | 4.6 (3.4, 6.0) | 6.3 (4.5, 8.4) | 5.8 (4.1, 7.9) | 2.7 (1.8, 3.9) | 2.2 (0.9, 3.3) | 0.6 (0.3, 0.8) | 1.6 (0.5, 2.9) | 0.7 (0.6, 0.9) |
| South Asia | 23.4 (15.9, 34.8) | 12.4 (5.2, 20.5) | 10.1 (4.1, 16.7) | 17.1 (7.5, 27.8) | 5.3 (1.8, 8.9) | 1.8 (0.9, 2.9) | 1.4 (0.6, 2.7) | 3.2 (1.4, 5.0) | 3.4 (1.2, 5.7) | 1.3 (0.5, 2.0) | 1.0 (0.0, 3.0) | 2.3 (1.5, 3.3) |
| Southeast Asia and Oceania | 31.7 (24.7, 40.5) | 6.1 (4.3, 8.6) | 10.0 (7.7, 12.8) | 5.6 (2.6, 8.1) | 2.2 (1.4, 3.0) | 1.3 (0.9, 1.8) | 0.3 (0.2, 0.5) | 2.4 (1.6, 3.4) | 2.7 (1.4, 4.0) | 0.4 (0.1, 0.7) | 0.0 (0.0, 0.5) | 0.5 (0.4, 0.7) |
| East Asia | 26.9 (17.2, 40.3) | 0.0 (0.0, 0.0) | 12.4 (7.9, 18.5) | 2.2 (1.2, 3.5) | 1.3 (0.8, 1.9) | 0.1 (0.0, 0.1) | 0.0 (0.0, 0.0) | 1.5 (0.9, 2.2) | 0.7 (0.4, 1.1) | 1.1 (0.5, 1.8) | 0.1 (0.0, 0.3) | 0.0 (0.0, 0.0) |
| **2019-2020** | | | | | | | | | | | | |
| Overall | 32.4 (23.1, 42.8) | 8.7 (6.7, 10.7) | 6.7 (4.7, 8.5) | 11.9 (5.9, 17.5) | 3.8 (2.1, 5.1) | 3.3 (2.5, 4.0) | 1.9 (1.4, 2.6) | 1.7 (1.3, 2.1) | 1.3 (0.9, 1.8) | 0.6 (0.3, 0.9) | 1.1 (0.4, 1.9) | 0.7 (0.5, 1.0) |
| Central America | 21.0 (15.1, 28.8) | 12.0 (7.9, 17.1) | 22.5 (12.8, 32.2) | 8.7 (3.6, 13.9) | 4.0 (2.0, 5.8) | 1.7 (1.0, 2.5) | 0.4 (0.1, 0.8) | 3.7 (2.0, 5.7) | 1.7 (0.7, 2.7) | 0.2 (0.0, 0.3) | 0.2 (0.0, 0.6) | 0.0 (0.0, 0.0) |
| South America | 13.0 (9.6, 17.4) | 13.9 (9.9, 19.0) | 19.4 (14.5, 25.4) | 6.5 (4.1, 9.1) | 5.1 (3.4, 7.2) | 3.3 (2.3, 4.7) | 0.4 (0.3, 0.5) | 1.3 (0.8, 1.7) | 1.9 (1.2, 2.8) | 0.3 (0.1, 0.5) | 0.2 (0.1, 0.4) | 0.0 (0.0, 0.0) |
| Eastern Europe | 23.5 (14.4, 36.6) | 0.8 (0.5, 1.2) | 12.1 (7.8, 18.5) | 8.1 (3.4, 14.0) | 1.7 (1.0, 2.6) | 0.1 (0.1, 0.2) | 0.0 (0.0, 0.1) | 0.1 (0.0, 0.1) | 6.3 (2.8, 11.0) | 1.6 (0.5, 2.9) | 0.3 (0.1, 0.6) | 0.1 (0.1, 0.2) |
| Central and Western Asia | 22.0 (13.6, 32.0) | 13.3 (8.7, 19.5) | 11.4 (7.3, 16.5) | 6.5 (2.8, 10.2) | 5.5 (3.1, 8.3) | 0.8 (0.5, 1.2) | 1.8 (0.7, 2.9) | 2.6 (1.6, 4.1) | 0.9 (0.4, 1.4) | 0.2 (0.0, 0.4) | 1.5 (0.0, 3.0) | 0.0 (0.0, 0.0) |
| West Africa | 45.3 (28.1, 64.9) | 5.6 (2.9, 8.2) | 6.2 (2.8, 8.9) | 13.9 (2.9, 24.0) | 3.8 (0.8, 5.8) | 2.2 (1.2, 3.2) | 1.5 (0.9, 2.6) | 1.2 (0.6, 1.7) | 0.8 (0.5, 1.2) | 0.6 (0.2, 1.0) | 0.9 (0.2, 2.1) | 0.0 (0.0, 0.0) |
| East and Southern Africa | 17.3 (14.2, 21.0) | 15.4 (11.0, 20.4) | 7.3 (4.3, 10.0) | 6.8 (3.3, 9.6) | 4.3 (2.9, 6.1) | 5.0 (3.6, 6.8) | 3.5 (2.3, 4.8) | 3.1 (1.8, 4.6) | 2.2 (0.7, 3.4) | 0.1 (0.0, 0.1) | 1.2 (0.3, 2.4) | 0.0 (0.0, 0.0) |
| South Asia | 13.0 (9.6, 17.6) | 12.5 (7.2, 18.5) | 6.3 (2.9, 10.0) | 12.8 (7.5, 19.8) | 4.1 (1.7, 6.4) | 5.2 (3.0, 7.4) | 1.9 (1.1, 2.9) | 2.1 (1.2, 3.0) | 2.0 (0.5, 3.8) | 1.0 (0.4, 1.6) | 1.6 (0.0, 4.0) | 3.6 (2.5, 4.9) |
| Southeast Asia and Oceania | 32.2 (23.5, 44.2) | 2.9 (2.0, 4.5) | 8.2 (5.6, 11.4) | 9.6 (0.3, 15.3) | 1.5 (0.7, 2.3) | 0.8 (0.3, 1.5) | 0.2 (0.1, 0.3) | 0.6 (0.3, 1.0) | 1.4 (0.4, 3.2) | 0.9 (0.3, 1.5) | 0.0 (0.0, 1.3) | 0.0 (0.0, 0.0) |
| East Asia | 19.9 (12.8, 30.1) | 0.0 (0.0, 0.0) | 13.3 (8.5, 20.3) | 2.3 (1.1, 3.8) | 1.3 (0.8, 1.9) | 0.0 (0.0, 0.0) | 0.1 (0.1, 0.2) | 0.6 (0.4, 1.0) | 0.7 (0.4, 1.3) | 0.8 (0.2, 1.5) | 0.1 (0.0, 0.3) | 0.0 (0.0, 0.0) |
| **2021-2022** | | | | | | | | | | | | |
| Overall | 26.7 (20.7, 34.1) | 10.1 (7.7, 12.6) | 7.3 (5.0, 9.2) | 7.0 (4.1, 9.7) | 2.5 (1.7, 3.2) | 2.4 (1.9, 3.0) | 2.0 (1.3, 2.8) | 1.2 (0.9, 1.6) | 1.5 (1.0, 2.0) | 1.0 (0.6, 1.3) | 0.7 (0.3, 1.1) | 1.4 (1.1, 1.8) |
| Central America | 11.7 (8.8, 15.6) | 9.5 (6.4, 13.2) | 15.8 (7.4, 23.4) | 6.1 (2.8, 9.3) | 4.4 (2.2, 6.3) | 2.7 (1.6, 3.9) | 0.1 (0.0, 0.2) | 2.2 (1.5, 3.2) | 2.8 (1.1, 4.5) | 1.2 (0.1, 2.1) | 0.5 (0.0, 1.3) | 0.0 (0.0, 0.0) |
| South America | 8.7 (6.7, 11.3) | 12.1 (9.0, 16.2) | 21.4 (15.6, 28.7) | 6.1 (4.1, 8.5) | 3.2 (2.2, 4.3) | 3.1 (2.2, 4.2) | 0.2 (0.1, 0.3) | 2.1 (1.4, 3.0) | 3.6 (2.2, 5.1) | 0.3 (0.1, 0.4) | 0.1 (0.0, 0.2) | 0.0 (0.0, 0.0) |
| Eastern Europe | 23.7 (15.8, 35.2) | 1.1 (0.8, 1.7) | 12.3 (8.0, 18.1) | 6.3 (3.4, 10.0) | 0.6 (0.4, 1.0) | 0.7 (0.5, 1.1) | 0.1 (0.1, 0.2) | 1.1 (0.6, 1.8) | 3.0 (1.2, 5.1) | 0.3 (0.0, 0.5) | 0.3 (0.0, 0.6) | 0.0 (0.0, 0.0) |
| Central and Western Asia | 16.7 (10.9, 24.1) | 9.6 (6.3, 14.0) | 12.2 (7.5, 17.8) | 4.1 (1.6, 6.3) | 4.0 (2.6, 5.9) | 1.3 (0.8, 1.9) | 2.3 (1.1, 3.9) | 0.4 (0.3, 0.6) | 0.7 (0.3, 1.1) | 0.0 (0.0, 0.1) | 0.5 (0.0, 1.3) | 0.0 (0.0, 0.0) |
| West Africa | 36.0 (23.8, 50.7) | 4.7 (2.5, 6.9) | 7.1 (3.1, 10.5) | 9.3 (3.8, 14.5) | 2.1 (0.7, 3.3) | 1.4 (0.8, 2.1) | 1.9 (0.8, 3.5) | 1.2 (0.5, 1.8) | 0.3 (0.2, 0.4) | 0.3 (0.1, 0.4) | 0.5 (0.1, 1.2) | 0.1 (0.1, 0.1) |
| East and Southern Africa | 19.9 (16.5, 24.0) | 18.4 (13.1, 24.8) | 5.6 (3.6, 7.5) | 5.1 (3.0, 7.2) | 3.4 (2.2, 4.7) | 5.2 (3.8, 6.8) | 3.9 (2.6, 5.4) | 1.4 (0.9, 1.9) | 2.6 (1.4, 3.8) | 1.2 (0.2, 2.1) | 1.3 (0.4, 2.3) | 0.0 (0.0, 0.0) |
| South Asia | 15.0 (10.8, 20.9) | 16.8 (7.8, 25.5) | 5.7 (2.5, 9.4) | 4.8 (2.6, 7.4) | 2.6 (0.7, 4.2) | 2.5 (1.2, 3.6) | 1.0 (0.5, 1.7) | 1.1 (0.5, 1.7) | 2.3 (0.6, 4.0) | 2.0 (0.6, 3.1) | 0.6 (0.0, 1.5) | 3.7 (2.6, 5.4) |
| Southeast Asia and Oceania | 20.5 (14.7, 28.1) | 5.4 (3.2, 8.5) | 16.4 (10.3, 22.6) | 2.9 (0.9, 4.5) | 3.1 (1.7, 4.6) | 1.3 (0.7, 2.0) | 0.1 (0.1, 0.2) | 1.7 (1.1, 2.5) | 4.4 (2.0, 7.3) | 2.3 (1.0, 3.5) | 0.5 (0.0, 1.1) | 7.5 (4.9, 11.1) |
| East Asia | 15.0 (9.3, 23.0) | 0.0 (0.0, 0.0) | 9.4 (5.9, 14.7) | 5.4 (2.8, 8.9) | 0.5 (0.3, 0.8) | 0.0 (0.0, 0.0) | 0.1 (0.0, 0.1) | 0.8 (0.5, 1.4) | 0.4 (0.1, 0.7) | 2.5 (0.6, 4.5) | 0.1 (0.0, 0.3) | 0.0 (0.0, 0.0) |

Attributable fractions are expressed as a percent. tEPEC=typical enteropathogenic *E. coli*. ETEC=heat-stable enterotoxigenic *E. coli*

**S2 Table. Pathogen-specific attributable fractions with 95% confidence intervals of diarrhea hospitalizations in children aged <5 years both overall and by geographic grouping in countries participating in Global Pediatric Diarrhea Surveillance, limited to sites that had introduced rotavirus vaccine, by two-year time periods during 2017-2022 (Underlying data for Figure 4).**

| | **Rotavirus** | ***Shigella*** | **Norovirus** | **Adenovirus 40/41** | **Sapovirus** | **ETEC** | ***Cryptosporidium*** | **Astrovirus** | ***C. jejuni/coli*** | ***Salmonella*** | **tEPEC** | ***V. cholerae*** |
|---|---|---|---|---|---|---|---|---|---|---|---|---|
| **2017-2018** | | | | | | | | | | | | |
| Overall | 19.2 (16.2, 22.5) | 14.6 (11.2, 18.6) | 7.2 (5.1, 9.2) | 7.2 (4.7, 9.9) | 4.3 (3.2, 5.6) | 4.9 (3.6, 6.5) | 4.2 (2.9, 5.7) | 2.8 (2.0, 3.8) | 2.3 (1.3, 3.2) | 0.8 (0.5, 1.1) | 1.5 (0.5, 2.6) | 1.0 (0.7, 1.3) |
| Central America | 20.1 (14.4, 27.3) | 17.3 (10.2, 25.7) | 14.1 (7.1, 21.1) | 9.3 (4.1, 14.5) | 3.4 (1.5, 5.1) | 2.6 (1.2, 4.1) | 1.3 (0.6, 2.3) | 5.0 (2.5, 7.5) | 3.2 (0.3, 5.6) | 0.5 (0.2, 0.8) | 0.7 (0.0, 2.3) | 0.0 (0.0, 0.0) |
| South America | 13.0 (9.6, 17.4) | 12.2 (8.9, 16.2) | 23.8 (17.5, 31.5) | 5.4 (3.5, 7.7) | 4.2 (3.1, 5.6) | 2.6 (1.8, 3.4) | 0.6 (0.4, 0.8) | 2.1 (1.4, 2.9) | 2.8 (1.6, 4.1) | 0.8 (0.3, 1.3) | 0.2 (0.1, 0.4) | 0.0 (0.0, 0.0) |
| Eastern Europe | 18.7 (12.3, 27.4) | 6.7 (4.5, 9.6) | 9.5 (6.3, 14.0) | 6.9 (3.0, 10.8) | 2.9 (1.8, 4.4) | 1.3 (0.8, 2.0) | 1.0 (0.5, 1.6) | 0.5 (0.2, 0.7) | 4.2 (1.8, 7.4) | 2.3 (0.5, 3.8) | 0.1 (0.0, 0.2) | 0.0 (0.0, 0.0) |
| Central and Western Asia | 20.3 (14.2, 28.0) | 10.5 (7.4, 14.3) | 10.0 (7.1, 13.4) | 3.3 (1.5, 5.2) | 6.2 (4.4, 8.5) | 3.8 (2.7, 5.2) | 2.9 (1.6, 4.6) | 3.2 (2.1, 4.7) | 0.6 (0.3, 0.8) | 0.1 (0.0, 0.2) | 0.4 (0.1, 0.8) | 0.0 (0.0, 0.0) |
| West Africa | 19.9 (14.5, 26.1) | 9.4 (6.5, 12.7) | 3.4 (2.4, 4.6) | 3.8 (1.6, 6.2) | 2.7 (2.0, 3.7) | 2.9 (2.0, 4.1) | 3.3 (1.9, 4.8) | 0.7 (0.5, 1.0) | 1.8 (0.7, 2.9) | 0.9 (0.3, 1.8) | 2.1 (0.6, 3.9) | 0.0 (0.0, 0.0) |
| East and Southern Africa | 19.2 (15.3, 24.0) | 16.1 (11.4, 21.9) | 7.7 (4.8, 10.4) | 4.6 (2.7, 6.4) | 4.4 (3.0, 6.0) | 6.4 (4.3, 8.9) | 5.2 (3.3, 7.5) | 2.9 (1.8, 4.4) | 2.5 (0.9, 3.9) | 0.6 (0.3, 0.9) | 1.4 (0.1, 2.8) | 0.5 (0.4, 0.7) |
| South Asia | 18.5 (12.2, 27.2) | 14.7 (5.9, 24.5) | 7.8 (2.8, 13.1) | 18.7 (6.8, 31.7) | 5.6 (1.6, 9.4) | 1.9 (0.8, 3.1) | 1.9 (0.7, 3.5) | 4.1 (1.7, 6.6) | 2.1 (0.9, 3.8) | 1.3 (0.2, 2.1) | 1.5 (0.0, 4.4) | 3.4 (2.2, 5.1) |
| Southeast Asia and Oceania | 14.4 (9.5, 21.3) | 17.9 (11.8, 26.3) | 13.8 (5.0, 21.1) | 7.6 (0.0, 12.8) | 5.1 (2.7, 8.1) | 4.3 (2.5, 6.6) | 0.9 (0.5, 1.6) | 2.1 (1.2, 3.3) | 2.1 (0.2, 3.8) | 0.0 (0.0, 0.0) | 2.5 (0.0, 5.4) | 0.0 (0.0, 0.0) |
| **2019-2020** | | | | | | | | | | | | |
| Overall | 15.9 (13.8, 18.3) | 12.9 (10.1, 16.0) | 6.9 (5.0, 8.8) | 8.8 (6.3, 11.8) | 4.0 (2.8, 5.2) | 4.7 (3.6, 5.8) | 2.6 (2.0, 3.3) | 2.5 (1.8, 3.2) | 2.1 (1.2, 3.0) | 0.5 (0.3, 0.8) | 1.5 (0.6, 2.6) | 1.5 (1.1, 2.1) |
| Central America | 21.0 (15.0, 29.4) | 12.0 (7.8, 17.1) | 22.6 (12.7, 32.0) | 8.7 (3.5, 13.8) | 4.1 (2.0, 6.0) | 1.7 (1.0, 2.5) | 0.4 (0.1, 0.8) | 3.7 (2.0, 5.7) | 1.7 (0.7, 2.6) | 0.2 (0.0, 0.3) | 0.2 (0.0, 0.6) | 0.0 (0.0, 0.0) |
| South America | 13.1 (9.6, 17.3) | 13.8 (9.9, 19.0) | 19.3 (14.4, 25.4) | 6.5 (4.0, 9.1) | 5.2 (3.4, 7.3) | 3.4 (2.3, 4.7) | 0.4 (0.3, 0.5) | 1.3 (0.8, 1.7) | 1.9 (1.2, 2.8) | 0.3 (0.1, 0.5) | 0.2 (0.1, 0.4) | 0.0 (0.0, 0.0) |
| Eastern Europe | 12.1 (7.8, 18.0) | 5.7 (3.8, 8.2) | 10.7 (6.9, 15.8) | 0.9 (0.4, 1.4) | 2.0 (1.3, 3.0) | 1.0 (0.6, 1.7) | 0.3 (0.1, 0.5) | 0.3 (0.1, 0.5) | 5.3 (2.2, 9.0) | 1.4 (0.4, 2.6) | 1.0 (0.0, 2.1) | 0.9 (0.6, 1.4) |
| Central and Western Asia | 22.0 (13.7, 32.3) | 13.4 (8.7, 19.5) | 11.4 (7.3, 16.5) | 6.5 (2.7, 10.1) | 5.5 (3.1, 8.2) | 0.8 (0.5, 1.2) | 1.8 (0.7, 2.9) | 2.6 (1.6, 4.1) | 0.9 (0.4, 1.4) | 0.2 (0.0, 0.4) | 1.5 (0.0, 3.0) | 0.0 (0.0, 0.0) |
| West Africa | 19.4 (15.7, 23.5) | 8.0 (5.9, 10.1) | 6.5 (5.0, 8.2) | 3.7 (2.3, 5.2) | 2.7 (2.0, 3.5) | 2.8 (2.1, 3.5) | 2.4 (1.6, 3.3) | 2.1 (1.5, 2.8) | 2.0 (1.3, 2.8) | 0.6 (0.3, 0.9) | 2.0 (0.7, 3.3) | 0.1 (0.1, 0.2) |
| East and Southern Africa | 17.3 (14.1, 21.0) | 15.4 (11.1, 20.3) | 7.4 (4.2, 10.0) | 6.8 (3.3, 9.6) | 4.3 (2.9, 6.2) | 5.0 (3.7, 6.8) | 3.5 (2.3, 4.7) | 3.1 (1.8, 4.6) | 2.2 (0.7, 3.4) | 0.1 (0.0, 0.1) | 1.2 (0.3, 2.4) | 0.0 (0.0, 0.0) |
| South Asia | 13.1 (9.5, 17.7) | 12.5 (7.2, 18.4) | 6.3 (3.0, 10.0) | 12.8 (7.5, 19.7) | 4.1 (1.7, 6.4) | 5.2 (3.0, 7.4) | 1.9 (1.1, 2.9) | 2.1 (1.2, 3.0) | 2.0 (0.5, 3.8) | 1.0 (0.4, 1.6) | 1.6 (0.0, 4.0) | 3.5 (2.5, 4.9) |
| Southeast Asia and Oceania | 11.1 (6.6, 17.0) | 19.3 (12.0, 28.3) | 5.5 (1.0, 8.6) | 8.0 (0.0, 13.0) | 0.0 (0.0, 0.0) | 10.1 (6.2, 15.4) | 3.1 (1.2, 5.2) | 0.0 (0.0, 0.0) | 0.0 (0.0, 0.0) | 0.0 (0.0, 0.0) | 2.4 (0.0, 5.8) | 0.0 (0.0, 0.0) |
| **2021-2022** | | | | | | | | | | | | |
| Overall | 18.4 (15.8, 21.5) | 16.4 (11.8, 21.1) | 6.0 (4.4, 7.9) | 5.7 (4.2, 7.3) | 2.9 (1.9, 3.8) | 3.7 (2.9, 4.6) | 2.4 (1.8, 3.1) | 1.2 (0.8, 1.5) | 2.4 (1.5, 3.3) | 1.5 (0.8, 2.2) | 1.0 (0.5, 1.6) | 1.8 (1.2, 2.5) |
| Central America | 11.8 (8.8, 15.6) | 9.5 (6.3, 13.2) | 15.9 (7.2, 23.4) | 6.1 (2.8, 9.3) | 4.4 (2.2, 6.3) | 2.7 (1.6, 3.9) | 0.1 (0.0, 0.2) | 2.2 (1.4, 3.3) | 2.8 (1.1, 4.4) | 1.2 (0.1, 2.1) | 0.5 (0.0, 1.3) | 0.0 (0.0, 0.0) |
| South America | 8.7 (6.8, 11.4) | 12.1 (9.0, 16.3) | 21.4 (15.5, 28.8) | 6.1 (4.1, 8.5) | 3.2 (2.3, 4.4) | 3.1 (2.2, 4.2) | 0.2 (0.1, 0.3) | 2.1 (1.4, 3.0) | 3.6 (2.2, 5.1) | 0.3 (0.1, 0.4) | 0.1 (0.0, 0.2) | 0.0 (0.0, 0.0) |
| Eastern Europe | 23.9 (15.7, 35.2) | 1.1 (0.8, 1.7) | 12.3 (8.0, 18.2) | 6.4 (3.4, 9.9) | 0.6 (0.4, 1.0) | 0.7 (0.5, 1.1) | 0.1 (0.1, 0.2) | 1.1 (0.6, 1.8) | 2.9 (1.2, 5.1) | 0.3 (0.0, 0.5) | 0.3 (0.0, 0.6) | 0.0 (0.0, 0.0) |
| Central and Western Asia | 16.7 (11.1, 24.0) | 9.6 (6.2, 14.0) | 12.2 (7.6, 17.9) | 4.1 (1.6, 6.2) | 4.0 (2.6, 5.9) | 1.3 (0.8, 1.9) | 2.3 (1.1, 4.0) | 0.4 (0.3, 0.6) | 0.7 (0.3, 1.1) | 0.0 (0.0, 0.1) | 0.5 (0.0, 1.3) | 0.0 (0.0, 0.0) |
| West Africa | 26.0 (20.5, 32.5) | 7.6 (5.0, 10.1) | 8.3 (6.3, 10.7) | 12.0 (6.2, 17.7) | 2.3 (1.6, 3.2) | 3.0 (2.2, 4.0) | 2.5 (1.5, 3.6) | 0.7 (0.5, 1.0) | 2.2 (1.0, 3.3) | 1.2 (0.6, 1.8) | 1.4 (0.4, 2.6) | 0.8 (0.5, 1.1) |
| East and Southern Africa | 19.9 (16.5, 24.1) | 18.3 (13.0, 24.8) | 5.6 (3.7, 7.5) | 5.1 (3.0, 7.1) | 3.4 (2.2, 4.7) | 5.3 (3.9, 6.9) | 3.9 (2.6, 5.4) | 1.4 (0.9, 2.0) | 2.6 (1.3, 3.8) | 1.2 (0.2, 2.1) | 1.3 (0.4, 2.3) | 0.0 (0.0, 0.0) |
| South Asia | 15.0 (10.8, 20.9) | 16.9 (7.8, 25.5) | 5.6 (2.5, 9.4) | 4.7 (2.5, 7.3) | 2.6 (0.7, 4.2) | 2.5 (1.3, 3.7) | 1.0 (0.5, 1.7) | 1.1 (0.5, 1.7) | 2.3 (0.6, 4.0) | 2.0 (0.5, 3.2) | 0.6 (0.0, 1.5) | 3.7 (2.6, 5.3) |
| Southeast Asia and Oceania | 14.1 (9.1, 20.8) | 12.3 (8.2, 17.9) | 14.4 (5.2, 22.0) | 15.3 (1.4, 27.0) | 3.5 (1.6, 5.7) | 0.0 (0.0, 0.0) | 4.5 (2.1, 7.9) | 0.0 (0.0, 0.0) | 3.4 (1.0, 6.0) | 0.0 (0.0, 0.0) | 0.1 (0.0, 0.2) | 0.0 (0.0, 0.0) |

Attributable fractions are expressed as a percent. tEPEC=typical enteropathogenic *E. coli*. ETEC=heat-stable enterotoxigenic *E. coli*

**S3 Table. Pathogen-specific attributable fractions with 95% confidence intervals of diarrhea hospitalizations in children aged <5 years by Global Pediatric Diarrhea Surveillance site, 2021-2022 (Underlying data for Figure 5).**

| Geographic Grouping | Surveillance Site | Rotavirus | *Shigella* | Norovirus | Adenovirus 40/41 | Sapovirus | ETEC | *Cryptosporidium* | Astrovirus | *C. jejuni/coli* | *Salmonella* | tEPEC | *V. cholerae* |
|---|---|---|---|---|---|---|---|---|---|---|---|---|---|
| Central America | Honduras | 10.3 (8.8, 10.9) | 7.7 (4.7, 9.5) | 20.1 (7.8, 23.6) | 7.7 (3.2, 10.1) | 4.0 (0.9, 5.2) | 2.9 (1.5, 3.7) | 0.1 (0.1, 0.2) | 2.2 (1.3, 2.9) | 2.6 (0.0, 3.7) | 0.0 (0.0, 0.0) | 0.4 (0.0, 1.3) | 0.0 (0.0, 0.0) |
| | Nicaragua | 15.4 (13.5, 16.9) | 13.8 (10.0, 16.0) | 10.3 (8.1, 11.8) | 3.5 (0.7, 4.7) | 5.6 (4.4, 6.6) | 2.8 (1.8, 3.3) | 0.0 (0.0, 0.0) | 2.6 (1.6, 3.2) | 3.9 (1.5, 5.6) | 3.9 (0.7, 5.5) | 0.7 (0.0, 1.7) | 0.0 (0.0, 0.0) |
| South America | Bolivia | 11.7 (9.5, 12.8) | 15.7 (11.9, 18.0) | 23.7 (9.3, 26.5) | 5.4 (2.2, 7.6) | 3.7 (0.6, 4.5) | 6.0 (2.8, 7.3) | 0.6 (0.3, 0.9) | 1.4 (0.6, 1.9) | 6.8 (1.0, 10.4) | 0.3 (0.1, 0.6) | 0.1 (0.0, 0.2) | 0.0 (0.0, 0.0) |
| | Ecuador | 6.9 (5.5, 8.0) | 6.2 (5.1, 6.9) | 22.1 (17.5, 24.8) | 9.4 (4.5, 12.3) | 3.2 (2.4, 3.7) | 0.9 (0.7, 1.0) | 0.1 (0.1, 0.2) | 1.5 (0.9, 1.9) | 3.9 (1.0, 5.5) | 0.9 (0.1, 1.3) | 0.0 (0.0, 0.0) | 0.0 (0.0, 0.0) |
| | Peru | 7.7 (6.4, 8.7) | 12.9 (9.7, 14.3) | 23.1 (17.3, 26.3) | 6.4 (4.1, 7.8) | 3.2 (2.2, 3.9) | 3.0 (2.3, 3.4) | 0.1 (0.1, 0.2) | 2.6 (1.7, 3.2) | 2.6 (1.1, 3.8) | 0.0 (0.0, 0.0) | 0.1 (0.0, 0.2) | 0.0 (0.0, 0.0) |
| | Paraguay | 14.4 (10.8, 16.2) | 10.4 (8.9, 11.6) | 11.6 (8.3, 13.1) | 3.0 (1.1, 3.4) | 2.8 (2.0, 3.4) | 2.0 (1.5, 2.4) | 0.2 (0.1, 0.2) | 1.2 (0.8, 1.5) | 4.7 (1.7, 6.9) | 1.4 (0.0, 1.8) | 0.6 (0.0, 1.1) | 0.0 (0.0, 0.0) |
| Eastern Europe | Moldova | 24.6 (19.7, 26.4) | 1.2 (1.1, 1.2) | 12.6 (10.1, 13.9) | 6.6 (4.3, 8.1) | 0.6 (0.4, 0.8) | 0.7 (0.6, 0.9) | 0.1 (0.1, 0.2) | 1.2 (0.7, 1.5) | 3.1 (1.2, 4.5) | 0.3 (0.0, 0.5) | 0.3 (0.0, 0.6) | 0.0 (0.0, 0.0) |
| | Ukraine* | --- | --- | --- | --- | --- | --- | --- | --- | --- | --- | --- | --- |
| Central and Western Asia | Armenia | 13.8 (11.3, 15.9) | 2.3 (1.9, 2.6) | 8.7 (7.3, 10.1) | 4.9 (2.3, 6.0) | 2.6 (2.0, 2.9) | 1.3 (1.0, 1.5) | 0.0 (0.0, 0.0) | 4.9 (3.6, 6.2) | 4.3 (2.3, 6.4) | 1.0 (0.1, 1.4) | 0.3 (0.0, 0.6) | 0.0 (0.0, 0.0) |
| | Tajikistan | 17.1 (14.0, 19.8) | 10.2 (8.2, 11.7) | 12.8 (9.3, 15.0) | 4.3 (1.5, 5.2) | 4.2 (3.3, 5.0) | 1.3 (1.0, 1.6) | 2.4 (1.2, 3.6) | 0.2 (0.1, 0.3) | 0.6 (0.2, 0.8) | 0.0 (0.0, 0.0) | 0.6 (0.0, 1.2) | 0.0 (0.0, 0.0) |
| | Uzbekistan* | --- | --- | --- | --- | --- | --- | --- | --- | --- | --- | --- | --- |
| West Africa | Benin | 23.6 (19.8, 24.8) | 12.5 (4.7, 13.6) | 3.5 (1.4, 6.0) | 14.1 (4.7, 18.4) | 0.0 (0.0, 0.0) | 3.0 (1.3, 4.7) | 1.8 (0.7, 2.4) | 0.1 (0.0, 0.1) | 2.6 (0.3, 3.5) | 0.0 (0.0, 0.0) | 0.0 (0.0, 1.4) | 3.5 (3.4, 3.7) |
| | Burkina Faso | 20.4 (15.6, 23.1) | 4.8 (3.6, 5.6) | 7.9 (6.2, 8.8) | 9.1 (0.0, 12.2) | 3.1 (1.9, 3.8) | 3.1 (2.2, 3.8) | 3.4 (2.0, 4.8) | 1.0 (0.6, 1.3) | 3.0 (0.3, 4.3) | 0.8 (0.1, 1.2) | 1.7 (0.0, 3.7) | 0.0 (0.0, 0.0) |
| | Côte d'Ivoire* | --- | --- | --- | --- | --- | --- | --- | --- | --- | --- | --- | --- |
| | Ghana | 37.6 (31.3, 41.7) | 10.1 (3.6, 12.5) | 12.3 (9.5, 13.5) | 17.0 (1.2, 26.9) | 3.0 (2.0, 3.6) | 3.1 (2.3, 3.7) | 1.8 (0.7, 2.5) | 0.7 (0.4, 0.8) | 1.1 (0.3, 1.8) | 2.7 (1.2, 3.8) | 1.7 (0.0, 3.0) | 0.0 (0.0, 0.0) |
| | Nigeria | 38.2 (30.5, 42.0) | 4.4 (2.0, 5.5) | 7.4 (3.0, 8.6) | 9.1 (3.5, 13.1) | 2.2 (0.5, 2.9) | 1.3 (0.5, 1.8) | 1.8 (0.6, 3.3) | 1.3 (0.6, 1.6) | 0.1 (0.0, 0.1) | 0.2 (0.0, 0.3) | 0.4 (0.0, 1.0) | 0.0 (0.0, 0.0) |
| East and Southern Africa | Ethiopia | 11.2 (8.8, 12.3) | 22.6 (16.0, 26.1) | 5.8 (2.7, 6.8) | 4.0 (0.6, 6.1) | 4.3 (2.5, 5.1) | 4.4 (3.3, 5.5) | 2.7 (1.1, 4.0) | 1.7 (1.0, 2.1) | 2.1 (0.1, 3.3) | 0.2 (0.0, 0.4) | 1.0 (0.0, 2.1) | 0.0 (0.0, 0.0) |
| | Madagascar | 35.1 (27.9, 37.9) | 24.5 (16.9, 28.7) | 4.0 (1.2, 5.4) | 5.5 (0.3, 8.2) | 3.4 (0.7, 4.6) | 12.4 (7.2, 15.0) | 6.7 (3.3, 9.3) | 1.7 (0.7, 2.1) | 5.6 (2.0, 8.2) | 5.7 (0.0, 8.8) | 1.3 (0.0, 3.5) | 0.0 (0.0, 0.0) |
| | Mauritius | 10.2 (7.7, 12.2) | 2.9 (2.2, 3.4) | 14.9 (9.9, 17.0) | 16.6 (2.4, 25.8) | 3.8 (2.9, 4.5) | 0.4 (0.3, 0.5) | 0.3 (0.2, 0.5) | 0.9 (0.5, 1.2) | 7.4 (1.4, 10.1) | 2.8 (0.0, 4.4) | 0.8 (0.0, 1.4) | 0.0 (0.0, 0.0) |
| | Uganda | 28.3 (22.3, 31.1) | 5.4 (4.3, 6.2) | 6.4 (4.0, 7.7) | 3.1 (0.0, 5.3) | 1.7 (1.3, 2.1) | 2.7 (1.8, 3.5) | 7.7 (3.9, 10.0) | 0.1 (0.1, 0.1) | 1.6 (0.1, 3.0) | 1.1 (0.0, 2.5) | 2.4 (0.1, 4.7) | 0.0 (0.0, 0.0) |
| | Zambia | 47.7 (38.9, 50.6) | 6.5 (4.8, 8.1) | 7.9 (3.3, 15.1) | 15.9 (12.8, 24.5) | 1.3 (0.0, 2.9) | 3.3 (1.9, 4.5) | 2.1 (1.3, 2.4) | 1.7 (0.0, 2.6) | 1.5 (0.0, 2.8) | 1.4 (0.0, 1.7) | 0.0 (0.0, 3.4) | 0.0 (0.0, 0.0) |
| | Zimbabwe | 22.4 (18.3, 24.5) | 13.8 (9.4, 15.4) | 7.1 (5.5, 7.9) | 11.4 (0.4, 18.0) | 3.1 (2.1, 4.1) | 6.7 (4.6, 8.2) | 0.9 (0.5, 1.3) | 1.9 (1.2, 2.4) | 4.6 (2.1, 6.2) | 0.6 (0.3, 0.8) | 3.3 (0.0, 5.9) | 0.0 (0.0, 0.0) |
| South Asia | India #1 | 17.8 (14.6, 19.4) | 5.2 (1.8, 6.3) | 7.4 (2.2, 8.7) | 16.7 (7.0, 25.0) | 2.0 (0.0, 2.7) | 0.3 (0.1, 0.4) | 1.0 (0.5, 1.5) | 3.0 (0.9, 3.9) | 2.4 (0.4, 3.4) | 0.0 (0.0, 0.0) | 0.1 (0.0, 0.5) | 2.7 (2.6, 2.8) |
| | India #2* | --- | --- | --- | --- | --- | --- | --- | --- | --- | --- | --- | --- |
| | India #3 | 14.6 (10.9, 16.0) | 13.9 (5.0, 18.2) | 6.6 (2.3, 7.9) | 11.2 (5.3, 15.6) | 2.1 (0.3, 2.9) | 2.2 (0.7, 2.7) | 3.5 (1.7, 5.5) | 2.0 (0.6, 2.7) | 2.3 (0.3, 3.5) | 0.3 (0.1, 0.4) | 0.8 (0.0, 6.1) | 2.8 (2.2, 3.1) |
| | India #4 | 5.6 (4.2, 6.4) | 7.0 (3.4, 8.9) | 11.4 (4.1, 13.9) | 11.3 (1.5, 16.4) | 8.9 (1.1, 11.8) | 1.7 (0.7, 2.2) | 0.0 (0.0, 0.0) | 4.1 (1.5, 5.5) | 1.5 (0.4, 2.3) | 0.0 (0.0, 0.0) | 0.2 (0.0, 0.6) | 0.0 (0.0, 0.0) |
| | India #5 | 17.8 (15.6, 18.8) | 20.0 (7.1, 24.4) | 7.6 (3.0, 9.5) | 5.7 (3.1, 7.7) | 3.1 (0.2, 4.0) | 0.5 (0.2, 0.7) | 0.0 (0.0, 0.0) | 0.7 (0.2, 0.9) | 1.9 (0.4, 3.1) | 2.6 (0.8, 3.3) | 0.1 (0.0, 0.7) | 0.0 (0.0, 0.0) |
| | Pakistan #1 | 6.9 (5.3, 8.0) | 14.8 (10.5, 17.8) | 2.9 (0.9, 4.1) | 3.1 (0.0, 4.9) | 3.2 (0.0, 5.3) | 10.5 (4.5, 12.9) | 5.2 (2.6, 7.5) | 3.0 (0.3, 4.4) | 4.0 (0.0, 7.2) | 1.4 (0.0, 2.0) | 2.4 (0.0, 6.0) | 9.8 (8.3, 10.5) |
| | Pakistan #2 | 10.6 (7.9, 11.9) | 6.8 (4.9, 8.0) | 2.3 (0.3, 4.3) | 2.3 (0.2, 3.7) | 0.9 (0.0, 1.3) | 5.6 (2.5, 7.0) | 2.0 (0.9, 2.9) | 1.7 (0.1, 2.6) | 3.2 (0.0, 6.2) | 0.6 (0.0, 0.9) | 1.1 (0.0, 2.9) | 17.9 (16.1, 18.7) |
| Southeast Asia and Oceania | Fiji | 14.5 (10.9, 16.2) | 12.6 (10.9, 13.3) | 15.5 (5.6, 16.7) | 15.5 (1.7, 22.4) | 3.6 (1.7, 4.6) | 0.0 (0.0, 0.0) | 4.5 (2.8, 6.8) | 0.0 (0.0, 0.0) | 3.7 (1.1, 5.1) | 0.0 (0.0, 0.0) | 0.1 (0.0, 0.2) | 0.0 (0.0, 0.0) |
| | Indonesia* | --- | --- | --- | --- | --- | --- | --- | --- | --- | --- | --- | --- |
| | Lao PDR | 19.3 (16.1, 21.3) | 3.4 (2.5, 3.8) | 17.7 (15.1, 19.2) | 6.3 (1.2, 8.9) | 3.8 (2.5, 4.7) | 2.2 (1.6, 2.6) | 0.4 (0.2, 0.6) | 0.7 (0.4, 0.8) | 2.3 (1.0, 3.1) | 2.2 (0.9, 3.1) | 0.0 (0.0, 0.1) | 0.0 (0.0, 0.0) |
| | Myanmar* | --- | --- | --- | --- | --- | --- | --- | --- | --- | --- | --- | --- |
| | Philippines | 22.9 (17.0, 24.5) | 8.5 (5.8, 10.9) | 17.1 (8.1, 19.6) | 3.3 (0.0, 4.9) | 4.4 (2.3, 5.2) | 2.1 (1.1, 2.5) | 0.2 (0.1, 0.3) | 0.9 (0.5, 1.1) | 7.0 (3.2, 9.5) | 3.5 (1.7, 4.3) | 0.8 (0.0, 1.5) | 12.6 (11.3, 13.0) |
| | Viet Nam | 18.4 (13.7, 21.9) | 0.7 (0.6, 0.8) | 16.3 (13.2, 18.8) | 1.8 (0.5, 2.5) | 1.2 (0.8, 1.4) | 0.0 (0.0, 0.0) | 0.0 (0.0, 0.0) | 3.6 (2.1, 4.6) | 0.7 (0.2, 0.9) | 0.5 (0.0, 0.8) | 0.0 (0.0, 0.0) | 0.0 (0.0, 0.0) |
| East Asia | China #1 | 7.1 (5.3, 8.1) | 0.0 (0.0, 0.0) | 1.7 (1.4, 2.0) | 2.8 (1.5, 3.3) | 0.1 (0.0, 0.1) | 0.0 (0.0, 0.0) | 0.0 (0.0, 0.0) | 0.5 (0.3, 0.7) | 0.1 (0.0, 0.1) | 0.4 (0.0, 0.7) | 0.0 (0.0, 0.0) | 0.0 (0.0, 0.0) |
| | China #2 | 20.8 (15.1, 23.0) | 0.0 (0.0, 0.0) | 9.5 (7.7, 10.5) | 9.4 (2.3, 12.7) | 1.0 (0.8, 1.2) | 0.0 (0.0, 0.0) | 0.0 (0.0, 0.0) | 1.8 (1.1, 2.1) | 0.9 (0.2, 1.2) | 6.3 (1.0, 9.0) | 0.0 (0.0, 0.0) | 0.0 (0.0, 0.0) |
| | China #3 | 18.7 (15.2, 20.7) | 0.0 (0.0, 0.0) | 18.0 (14.5, 19.8) | 5.1 (2.9, 6.9) | 0.4 (0.3, 0.5) | 0.0 (0.0, 0.0) | 0.2 (0.1, 0.3) | 0.4 (0.2, 0.5) | 0.2 (0.1, 0.4) | 1.2 (0.3, 1.8) | 0.4 (0.0, 0.8) | 0.0 (0.0, 0.0) |

Attributable fractions are expressed as a percent. Lao PDR=Lao People's Democratic Republic. tEPEC=typical enteropathogenic *E. coli*. ETEC=enterotoxigenic *E. coli*.
*Site did not participate in GPDS during this two-year time period

**S4 Table. Pathogen-specific attributable fractions with 95% confidence intervals of diarrhea hospitalizations in children aged <5 years by Global Pediatric Diarrhea Surveillance site, 2019-2020 (Underlying data for Figure 5).**

| Geographic Grouping | Surveillance Site | Rotavirus | *Shigella* | Norovirus | Adenovirus 40/41 | Sapovirus | ETEC | *Cryptosporidium* | Astrovirus | *C. jejuni/coli* | *Salmonella* | tEPEC | *V. cholerae* |
|---|---|---|---|---|---|---|---|---|---|---|---|---|---|
| Central America | Honduras | 25.2 (22.2, 27.1) | 11.4 (6.7, 14.4) | 20.7 (5.6, 24.3) | 9.8 (3.2, 14.5) | 3.9 (0.9, 4.9) | 1.8 (0.7, 2.3) | 0.6 (0.3, 1.0) | 4.1 (1.9, 5.6) | 1.5 (0.0, 2.4) | 0.1 (0.0, 0.1) | 0.2 (0.0, 0.5) | 0.0 (0.0, 0.0) |
| | Nicaragua | 14.1 (11.2, 15.5) | 13.8 (10.7, 15.3) | 31.3 (25.7, 35.0) | 7.3 (1.8, 11.8) | 5.0 (3.4, 6.3) | 1.9 (1.4, 2.2) | 0.0 (0.0, 0.0) | 3.2 (1.6, 4.2) | 2.2 (0.7, 3.0) | 0.4 (0.0, 0.6) | 0.4 (0.0, 0.7) | 0.0 (0.0, 0.0) |
| South America | Bolivia | 17.1 (14.0, 18.6) | 15.6 (11.5, 18.1) | 26.2 (10.0, 30.4) | 9.9 (2.5, 13.9) | 3.3 (0.4, 4.2) | 6.1 (2.9, 7.6) | 0.4 (0.1, 0.6) | 2.5 (1.1, 3.3) | 2.4 (1.2, 3.3) | 0.7 (0.2, 0.9) | 0.2 (0.0, 0.7) | 0.0 (0.0, 0.0) |
| | Ecuador | 12.5 (9.8, 13.9) | 7.8 (6.5, 8.5) | 29.0 (23.2, 31.8) | 7.2 (2.4, 9.3) | 3.9 (3.0, 4.8) | 1.5 (1.1, 1.7) | 1.1 (0.6, 1.7) | 1.9 (1.1, 2.4) | 2.6 (1.0, 4.1) | 0.5 (0.1, 0.7) | 0.2 (0.0, 0.4) | 0.0 (0.0, 0.0) |
| | Peru | 13.5 (10.0, 14.9) | 15.9 (12.7, 18.1) | 17.9 (14.1, 20.2) | 5.8 (3.8, 7.5) | 6.1 (4.1, 7.4) | 3.6 (2.4, 4.3) | 0.2 (0.1, 0.3) | 1.1 (0.6, 1.4) | 1.5 (0.6, 2.4) | 0.2 (0.0, 0.4) | 0.2 (0.0, 0.4) | 0.0 (0.0, 0.0) |
| | Paraguay | 6.1 (4.9, 6.9) | 5.1 (3.9, 6.1) | 12.1 (8.8, 13.5) | 9.3 (3.1, 12.9) | 3.8 (2.7, 4.6) | 0.4 (0.3, 0.5) | 1.0 (0.4, 1.4) | 0.2 (0.1, 0.3) | 4.3 (1.4, 6.0) | 0.1 (0.0, 0.1) | 0.6 (0.0, 1.5) | 0.0 (0.0, 0.0) |
| Eastern Europe | Moldova | 12.6 (9.3, 13.8) | 5.8 (5.1, 6.4) | 11.0 (8.9, 12.4) | 0.9 (0.6, 1.2) | 2.1 (1.7, 2.4) | 1.0 (0.6, 1.4) | 0.3 (0.1, 0.4) | 0.3 (0.1, 0.4) | 5.5 (2.8, 7.7) | 1.5 (0.5, 2.2) | 1.0 (0.0, 1.9) | 1.0 (0.8, 1.0) |
| | Ukraine | 26.2 (21.4, 28.9) | 0.0 (0.0, 0.0) | 12.7 (11.0, 14.0) | 9.7 (4.6, 12.5) | 1.7 (1.1, 2.0) | 0.0 (0.0, 0.0) | 0.0 (0.0, 0.0) | 0.0 (0.0, 0.0) | 7.0 (2.9, 9.4) | 1.7 (0.4, 2.6) | 0.2 (0.0, 0.4) | 0.0 (0.0, 0.0) |
| Central and Western Asia | Armenia | 5.4 (4.2, 6.2) | 4.5 (4.0, 4.8) | 4.1 (3.4, 4.7) | 11.7 (5.1, 16.3) | 1.2 (0.8, 1.5) | 2.9 (2.0, 3.7) | 0.0 (0.0, 0.0) | 0.0 (0.0, 0.0) | 4.9 (2.7, 7.0) | 4.6 (0.7, 6.6) | 0.0 (0.0, 0.0) | 0.0 (0.0, 0.0) |
| | Tajikistan | 23.6 (17.6, 26.4) | 14.2 (11.0, 16.5) | 12.1 (8.9, 13.6) | 6.6 (2.8, 8.5) | 5.9 (3.5, 7.0) | 0.7 (0.5, 0.9) | 1.9 (0.8, 2.5) | 2.8 (2.1, 3.6) | 0.7 (0.2, 1.0) | 0.0 (0.0, 0.0) | 1.6 (0.0, 2.9) | 0.0 (0.0, 0.0) |
| | Uzbekistan* | --- | --- | --- | --- | --- | --- | --- | --- | --- | --- | --- | --- |
| West Africa | Benin | 36.9 (30.6, 38.6) | 19.2 (10.0, 20.1) | 3.8 (1.7, 6.8) | 7.3 (1.0, 10.9) | 2.1 (0.3, 3.4) | 3.8 (1.5, 5.5) | 3.5 (1.4, 4.4) | 1.6 (0.4, 2.1) | 4.7 (1.8, 5.9) | 0.4 (0.2, 0.5) | 0.0 (0.0, 3.2) | 0.9 (0.9, 0.9) |
| | Burkina Faso | 15.7 (11.9, 17.6) | 4.6 (3.7, 5.4) | 4.3 (3.5, 4.8) | 2.3 (1.2, 3.0) | 5.1 (3.9, 6.3) | 1.5 (1.2, 1.9) | 0.7 (0.4, 1.0) | 3.7 (2.4, 4.6) | 2.5 (1.0, 3.5) | 0.3 (0.0, 0.5) | 2.3 (0.0, 4.1) | 0.0 (0.0, 0.0) |
| | Côte d'Ivoire | 10.1 (7.4, 11.4) | 5.9 (4.4, 6.8) | 6.7 (4.4, 8.7) | 1.2 (0.0, 2.2) | 0.8 (0.6, 1.0) | 2.2 (1.6, 2.9) | 3.3 (1.5, 4.7) | 2.0 (1.0, 2.6) | 1.0 (0.2, 1.8) | 0.6 (0.0, 1.4) | 3.2 (0.0, 5.8) | 0.0 (0.0, 0.0) |
| | Ghana | 26.0 (22.7, 28.2) | 9.1 (3.6, 11.9) | 10.9 (8.2, 12.2) | 6.9 (2.4, 10.1) | 2.4 (1.8, 2.8) | 4.5 (3.1, 5.2) | 2.9 (1.5, 4.3) | 0.6 (0.5, 0.9) | 1.5 (0.3, 2.3) | 1.0 (0.3, 1.3) | 1.2 (0.0, 2.2) | 0.0 (0.0, 0.0) |
| | Nigeria | 51.9 (37.3, 57.0) | 5.4 (2.6, 6.8) | 6.5 (2.5, 7.5) | 16.1 (3.7, 23.0) | 4.1 (0.6, 5.5) | 2.3 (1.0, 2.7) | 1.4 (0.6, 2.3) | 1.0 (0.4, 1.3) | 0.6 (0.2, 0.8) | 0.7 (0.2, 0.9) | 0.7 (0.0, 1.9) | 0.0 (0.0, 0.0) |
| East and Southern Africa | Ethiopia | 11.0 (9.0, 13.0) | 17.8 (11.7, 20.2) | 9.0 (3.9, 10.6) | 5.9 (0.0, 8.7) | 6.2 (4.1, 7.3) | 5.7 (4.1, 6.8) | 2.1 (0.9, 3.3) | 4.8 (2.6, 5.9) | 2.6 (0.0, 3.9) | 0.0 (0.0, 0.0) | 1.3 (0.0, 3.0) | 0.0 (0.0, 0.0) |
| | Madagascar | 23.3 (17.7, 25.7) | 17.9 (10.9, 21.1) | 6.4 (1.9, 8.4) | 9.9 (1.7, 14.7) | 1.8 (0.6, 2.6) | 5.8 (2.9, 7.5) | 4.9 (1.9, 6.5) | 1.0 (0.6, 1.3) | 1.3 (0.4, 1.8) | 0.1 (0.0, 0.3) | 1.2 (0.0, 2.6) | 0.0 (0.0, 0.0) |
| | Mauritius | 6.7 (5.0, 7.6) | 1.4 (1.2, 1.5) | 7.0 (4.4, 7.8) | 9.9 (1.2, 13.7) | 4.5 (3.3, 5.3) | 0.2 (0.1, 0.3) | 0.0 (0.0, 0.0) | 0.0 (0.0, 0.0) | 4.1 (1.1, 6.3) | 1.9 (0.0, 2.8) | 1.4 (0.0, 2.6) | 0.0 (0.0, 0.0) |
| | Uganda | 17.8 (13.6, 19.9) | 5.1 (3.8, 6.0) | 6.3 (3.9, 7.7) | 3.9 (0.0, 6.5) | 1.8 (1.5, 2.2) | 4.2 (2.9, 5.2) | 7.3 (3.3, 10.1) | 1.0 (0.6, 1.4) | 1.9 (0.2, 3.8) | 0.0 (0.0, 0.0) | 1.3 (0.0, 2.6) | 0.0 (0.0, 0.0) |
| | Zambia | 48.7 (38.7, 52.0) | 24.2 (18.6, 28.4) | 6.0 (2.6, 12.1) | 19.9 (14.6, 30.9) | 2.5 (0.0, 7.4) | 4.5 (2.3, 6.2) | 4.6 (2.0, 5.5) | 1.4 (0.0, 2.4) | 1.8 (0.0, 3.1) | 0.0 (0.0, 0.0) | 0.0 (0.0, 5.6) | 0.0 (0.0, 0.0) |
| | Zimbabwe | 34.6 (27.3, 38.2) | 11.6 (7.0, 13.7) | 0.8 (0.7, 0.9) | 3.6 (0.2, 5.1) | 2.5 (1.8, 3.1) | 1.8 (1.4, 2.2) | 2.0 (1.0, 2.8) | 1.5 (1.1, 2.0) | 3.7 (1.5, 5.4) | 1.7 (0.4, 2.3) | 2.2 (0.0, 3.9) | 0.0 (0.0, 0.0) |
| South Asia | India #1 | 17.7 (14.3, 19.0) | 20.5 (9.0, 24.9) | 7.2 (2.4, 8.8) | 25.6 (13.8, 36.0) | 3.2 (0.2, 4.3) | 3.4 (1.6, 4.1) | 0.5 (0.2, 0.7) | 3.0 (1.3, 3.8) | 1.6 (0.1, 2.4) | 1.2 (0.1, 1.6) | 1.0 (0.0, 2.4) | 1.4 (0.9, 1.6) |
| | India #2* | --- | --- | --- | --- | --- | --- | --- | --- | --- | --- | --- | --- |
| | India #3 | 12.3 (10.5, 13.6) | 11.6 (4.9, 15.6) | 8.1 (2.4, 9.5) | 18.8 (2.8, 27.7) | 6.7 (1.0, 9.3) | 2.9 (1.3, 3.5) | 2.0 (1.0, 3.2) | 2.9 (1.6, 3.9) | 1.8 (0.3, 2.5) | 0.6 (0.2, 0.8) | 0.7 (0.0, 4.5) | 0.4 (0.3, 0.4) |
| | India #4 | 10.8 (8.2, 12.3) | 7.2 (2.6, 9.6) | 10.0 (3.5, 12.3) | 6.7 (2.1, 10.4) | 3.9 (0.7, 5.5) | 3.1 (1.2, 4.3) | 1.3 (0.5, 2.2) | 1.8 (0.5, 2.3) | 0.9 (0.1, 1.4) | 0.9 (0.1, 1.3) | 0.6 (0.0, 3.0) | 0.0 (0.0, 0.0) |
| | India #5* | --- | --- | --- | --- | --- | --- | --- | --- | --- | --- | --- | --- |
| | Pakistan #1 | 12.9 (10.1, 14.2) | 12.7 (9.1, 15.0) | 4.0 (0.8, 6.3) | 5.3 (0.0, 8.4) | 4.0 (0.2, 5.8) | 10.6 (4.0, 13.0) | 3.3 (1.4, 4.8) | 1.2 (0.3, 1.9) | 3.9 (0.0, 7.7) | 1.4 (0.0, 2.2) | 2.7 (0.0, 8.1) | 10.4 (9.5, 10.8) |
| | Pakistan #2* | --- | --- | --- | --- | --- | --- | --- | --- | --- | --- | --- | --- |
| Southeast Asia and Oceania | Fiji | 11.6 (7.5, 13.4) | 19.7 (14.6, 21.8) | 5.9 (1.2, 6.6) | 8.6 (0.0, 10.2) | 0.0 (0.0, 0.0) | 10.5 (7.2, 12.1) | 3.1 (1.5, 4.5) | 0.0 (0.0, 0.0) | 0.0 (0.0, 0.0) | 0.0 (0.0, 0.0) | 2.4 (0.0, 5.2) | 0.0 (0.0, 0.0) |
| | Indonesia* | --- | --- | --- | --- | --- | --- | --- | --- | --- | --- | --- | --- |
| | Lao PDR | 46.0 (34.9, 51.5) | 4.5 (3.3, 4.8) | 11.4 (9.4, 12.8) | 2.8 (1.0, 3.7) | 1.9 (1.2, 2.4) | 1.5 (1.2, 1.8) | 0.7 (0.4, 1.1) | 0.0 (0.0, 0.0) | 3.0 (1.0, 4.2) | 4.4 (0.9, 5.7) | 0.2 (0.0, 0.3) | 0.0 (0.0, 0.0) |
| | Myanmar | 60.4 (53.9, 63.6) | 5.7 (4.5, 8.4) | 7.4 (3.9, 10.8) | 23.6 (1.3, 29.5) | 1.8 (0.0, 2.8) | 1.7 (0.6, 2.7) | 0.4 (0.3, 0.6) | 0.7 (0.0, 1.1) | 1.8 (0.0, 4.9) | 1.4 (0.0, 1.7) | 0.0 (0.0, 2.7) | 0.0 (0.0, 0.0) |
| | Philippines* | --- | --- | --- | --- | --- | --- | --- | --- | --- | --- | --- | --- |
| | Viet Nam | 8.2 (6.2, 10.0) | 0.4 (0.3, 0.5) | 8.9 (5.8, 10.5) | 0.1 (0.0, 0.2) | 1.3 (0.9, 1.6) | 0.0 (0.0, 0.0) | 0.0 (0.0, 0.0) | 0.7 (0.4, 0.8) | 1.1 (0.3, 1.5) | 0.5 (0.0, 0.9) | 0.0 (0.0, 0.1) | 0.0 (0.0, 0.0) |
| East Asia | China #1 | 9.1 (7.3, 10.6) | 0.0 (0.0, 0.0) | 7.3 (6.0, 8.1) | 0.6 (0.3, 0.9) | 0.9 (0.6, 1.1) | 0.0 (0.0, 0.0) | 0.0 (0.0, 0.0) | 0.5 (0.3, 0.6) | 0.6 (0.2, 0.9) | 0.2 (0.0, 0.3) | 0.0 (0.0, 0.0) | 0.0 (0.0, 0.0) |
| | China #2 | 13.6 (9.9, 15.0) | 0.0 (0.0, 0.0) | 11.5 (9.8, 12.3) | 2.6 (1.2, 3.4) | 0.5 (0.3, 0.6) | 0.0 (0.0, 0.0) | 0.0 (0.0, 0.0) | 1.1 (0.7, 1.3) | 1.7 (0.6, 2.5) | 2.0 (0.2, 3.1) | 0.2 (0.0, 0.4) | 0.0 (0.0, 0.0) |
| | China #3 | 38.9 (31.8, 43.3) | 0.0 (0.0, 0.0) | 22.4 (18.4, 24.6) | 4.1 (1.6, 5.6) | 2.5 (1.8, 3.0) | 0.0 (0.0, 0.0) | 0.4 (0.2, 0.5) | 0.5 (0.3, 0.6) | 0.0 (0.0, 0.0) | 0.3 (0.0, 0.5) | 0.3 (0.0, 0.5) | 0.0 (0.0, 0.0) |

Attributable fractions are expressed as a percent. Lao PDR=Lao People's Democratic Republic. tEPEC=typical enteropathogenic *E. coli*. ETEC=enterotoxigenic *E. coli*.

*Site did not participate in GPDS during this two-year time period

**S5 Table. Pathogen-specific attributable fractions with 95% confidence intervals of diarrhea hospitalizations in children aged <5 years by Global Pediatric Diarrhea Surveillance site, 2017-2018 (Underlying data for Figure 5).**

| Geographic Grouping | Surveillance Site | Rotavirus | *Shigella* | Norovirus | Adenovirus 40/41 | Sapovirus | ETEC | *Cryptosporidium* | Astrovirus | *C. jejuni/coli* | *Salmonella* | tEPEC | *V. cholerae* |
|---|---|---|---|---|---|---|---|---|---|---|---|---|---|
| Central America | Honduras | 23.2 (20.3, 25.2) | 20.8 (12.1, 26.4) | 16.7 (5.9, 20.1) | 11.7 (4.3, 16.1) | 3.3 (0.5, 4.4) | 3.7 (1.8, 4.5) | 1.7 (0.7, 2.6) | 5.5 (2.2, 7.5) | 4.7 (0.3, 7.1) | 0.4 (0.1, 0.7) | 1.0 (0.0, 3.0) | 0.0 (0.0, 0.0) |
| | Nicaragua | 15.0 (11.4, 16.9) | 10.9 (7.2, 13.2) | 12.7 (9.8, 14.4) | 5.0 (1.0, 7.1) | 4.1 (2.9, 5.0) | 0.8 (0.6, 1.0) | 0.6 (0.3, 0.9) | 4.8 (2.8, 6.0) | 0.6 (0.2, 0.8) | 0.7 (0.2, 0.9) | 0.3 (0.0, 0.4) | 0.0 (0.0, 0.0) |
| South America | Bolivia | 19.9 (15.6, 22.6) | 12.1 (9.2, 14.2) | 24.1 (9.9, 27.4) | 1.7 (0.4, 3.0) | 3.9 (0.8, 4.7) | 7.0 (2.3, 8.2) | 1.6 (0.6, 2.6) | 1.8 (0.8, 2.4) | 3.8 (1.7, 5.3) | 0.0 (0.0, 0.0) | 0.2 (0.0, 0.5) | 0.0 (0.0, 0.0) |
| | Ecuador | 7.5 (5.8, 8.4) | 5.4 (4.5, 5.8) | 25.3 (19.5, 27.8) | 6.1 (1.7, 8.5) | 4.8 (3.5, 5.6) | 2.7 (2.1, 3.2) | 0.7 (0.4, 0.9) | 3.1 (2.1, 3.8) | 3.4 (1.6, 4.9) | 1.2 (0.2, 1.8) | 0.4 (0.0, 0.7) | 0.0 (0.0, 0.0) |
| | Peru | 13.6 (9.8, 15.4) | 13.8 (11.6, 15.3) | 25.1 (19.6, 27.9) | 5.8 (3.5, 7.6) | 4.6 (3.7, 5.1) | 1.8 (1.3, 2.1) | 0.3 (0.2, 0.4) | 2.2 (1.3, 2.7) | 2.7 (1.2, 3.7) | 1.0 (0.3, 1.6) | 0.2 (0.0, 0.3) | 0.0 (0.0, 0.0) |
| | Paraguay | 7.5 (4.9, 8.4) | 13.6 (10.4, 15.0) | 15.8 (12.1, 18.0) | 11.8 (3.4, 15.7) | 2.0 (1.7, 2.3) | 1.3 (0.9, 1.7) | 0.8 (0.4, 1.2) | 1.0 (0.6, 1.3) | 1.4 (0.7, 1.8) | 0.0 (0.0, 0.0) | 0.8 (0.0, 1.4) | 0.0 (0.0, 0.0) |
| Eastern Europe | Moldova | 19.4 (15.3, 21.0) | 6.9 (5.9, 7.4) | 9.6 (7.9, 10.8) | 7.3 (3.3, 8.7) | 3.0 (2.2, 3.5) | 1.4 (1.1, 1.6) | 1.0 (0.5, 1.4) | 0.5 (0.3, 0.6) | 4.4 (2.1, 6.5) | 2.4 (0.6, 3.2) | 0.1 (0.0, 0.2) | 0.0 (0.0, 0.0) |
| | Ukraine | 36.9 (30.3, 40.0) | 0.3 (0.3, 0.4) | 7.8 (6.7, 8.5) | 1.3 (0.6, 1.9) | 3.0 (2.1, 3.5) | 0.8 (0.6, 0.9) | 0.0 (0.0, 0.0) | 0.8 (0.4, 0.9) | 4.7 (2.1, 6.5) | 1.7 (0.2, 2.5) | 0.4 (0.0, 0.9) | 0.0 (0.0, 0.0) |
| Central and Western Asia | Armenia | 11.9 (9.7, 13.4) | 8.7 (7.7, 9.5) | 8.5 (6.3, 9.5) | 2.0 (1.0, 2.3) | 1.7 (1.1, 2.0) | 3.1 (2.2, 3.7) | 0.2 (0.2, 0.3) | 1.6 (1.0, 2.0) | 6.0 (2.4, 8.3) | 0.5 (0.1, 0.7) | 0.0 (0.0, 0.1) | 0.0 (0.0, 0.0) |
| | Tajikistan | 25.5 (19.7, 28.6) | 12.5 (10.2, 14.3) | 8.0 (5.5, 9.1) | 3.6 (1.5, 5.1) | 6.9 (5.4, 8.0) | 3.9 (3.1, 4.7) | 4.0 (2.2, 5.7) | 4.3 (3.1, 5.3) | 0.4 (0.1, 0.6) | 0.0 (0.0, 0.0) | 0.4 (0.0, 0.9) | 0.0 (0.0, 0.0) |
| | Uzbekistan | 10.2 (7.5, 11.1) | 6.2 (5.0, 7.0) | 15.7 (11.2, 19.0) | 3.1 (0.0, 5.2) | 5.3 (4.1, 6.2) | 3.8 (2.7, 4.7) | 0.9 (0.4, 1.3) | 1.2 (0.8, 1.7) | 0.2 (0.0, 0.4) | 0.2 (0.0, 0.6) | 0.4 (0.0, 1.1) | 0.0 (0.0, 0.0) |
| West Africa | Benin | 35.9 (31.4, 38.0) | 18.7 (10.1, 19.4) | 3.2 (1.4, 5.8) | 12.4 (5.2, 16.6) | 1.7 (0.2, 2.6) | 1.7 (0.9, 2.3) | 4.2 (2.1, 5.2) | 1.5 (0.4, 2.4) | 3.2 (1.1, 4.4) | 0.9 (0.5, 1.1) | 0.0 (0.0, 6.8) | 0.1 (0.1, 0.1) |
| | Burkina Faso | 33.3 (23.7, 38.0) | 9.6 (7.3, 10.8) | 5.8 (4.3, 7.0) | 4.2 (1.5, 5.7) | 4.3 (2.8, 5.5) | 0.8 (0.6, 1.0) | 3.7 (1.8, 5.1) | 2.7 (1.8, 3.3) | 1.1 (0.4, 1.7) | 0.5 (0.0, 0.7) | 0.6 (0.0, 1.3) | 0.0 (0.0, 0.0) |
| | Côte d'Ivoire | 9.3 (7.4, 10.4) | 10.7 (7.8, 12.2) | 1.1 (0.7, 1.5) | 3.3 (0.3, 5.7) | 2.9 (2.3, 3.4) | 3.6 (2.3, 4.5) | 2.2 (1.4, 3.0) | 0.6 (0.3, 0.8) | 1.2 (0.1, 2.3) | 0.6 (0.0, 1.8) | 2.2 (0.0, 4.6) | 0.0 (0.0, 0.0) |
| | Ghana | 35.6 (27.2, 39.0) | 8.2 (3.6, 10.0) | 6.6 (5.5, 7.6) | 4.5 (1.2, 7.2) | 2.6 (1.6, 3.4) | 2.1 (1.4, 2.6) | 5.0 (1.7, 6.9) | 0.9 (0.6, 1.1) | 2.5 (0.7, 3.8) | 1.3 (0.6, 1.9) | 2.1 (0.0, 3.5) | 0.0 (0.0, 0.0) |
| | Nigeria | 52.7 (45.7, 58.8) | 9.6 (4.8, 12.7) | 4.1 (1.6, 4.9) | 5.5 (1.8, 7.2) | 4.9 (1.6, 6.5) | 6.6 (2.9, 7.9) | 3.4 (1.5, 5.5) | 4.0 (2.2, 5.1) | 0.1 (0.0, 0.2) | 2.3 (0.5, 3.2) | 0.2 (0.0, 0.5) | 0.1 (0.1, 0.1) |
| East and Southern Africa | Ethiopia | 12.9 (10.6, 15.1) | 17.8 (14.0, 20.4) | 6.5 (2.6, 7.7) | 1.8 (0.0, 2.7) | 4.6 (2.9, 5.7) | 6.9 (4.2, 8.5) | 5.2 (2.7, 7.4) | 3.9 (2.5, 4.9) | 2.6 (0.0, 3.6) | 0.0 (0.0, 0.0) | 1.7 (0.0, 3.2) | 0.7 (0.6, 0.7) |
| | Madagascar | 28.4 (22.4, 30.8) | 14.7 (9.8, 17.4) | 11.5 (4.6, 14.1) | 8.3 (0.0, 13.3) | 4.6 (1.8, 5.9) | 6.2 (2.0, 7.6) | 3.2 (1.3, 4.6) | 1.3 (0.7, 1.7) | 3.3 (1.1, 4.6) | 1.3 (0.0, 1.7) | 1.4 (0.0, 3.1) | 0.0 (0.0, 0.0) |
| | Mauritius | 11.5 (9.1, 12.9) | 1.9 (1.5, 2.2) | 18.0 (11.9, 20.0) | 12.7 (1.2, 18.4) | 6.3 (4.5, 7.8) | 1.5 (1.0, 1.9) | 0.2 (0.1, 0.3) | 3.1 (2.1, 3.8) | 2.0 (0.4, 3.1) | 2.6 (0.0, 3.8) | 0.9 (0.0, 2.4) | 0.0 (0.0, 0.0) |
| | Uganda | 26.5 (20.3, 30.5) | 6.1 (4.8, 7.1) | 4.5 (3.0, 5.6) | 5.8 (0.1, 9.5) | 5.6 (4.5, 6.8) | 5.5 (4.1, 6.9) | 9.2 (5.2, 11.9) | 1.7 (0.8, 2.3) | 0.9 (0.1, 1.5) | 0.4 (0.0, 1.1) | 2.6 (0.0, 5.0) | 1.7 (1.6, 1.8) |
| | Zambia | 46.2 (35.3, 48.7) | 13.5 (10.3, 16.6) | 10.0 (3.3, 19.9) | 20.4 (15.5, 28.6) | 3.5 (0.0, 8.7) | 7.1 (4.2, 9.4) | 13.2 (5.9, 15.2) | 1.3 (0.1, 2.5) | 2.7 (0.0, 4.9) | 2.3 (0.5, 2.7) | 0.0 (0.0, 6.0) | 1.0 (0.9, 1.0) |
| | Zimbabwe | 29.8 (22.3, 32.8) | 9.2 (6.0, 10.9) | 10.7 (8.7, 12.4) | 4.0 (0.0, 6.8) | 4.7 (3.1, 6.1) | 3.1 (2.0, 3.9) | 2.2 (1.3, 3.1) | 0.6 (0.3, 0.8) | 1.8 (0.5, 2.4) | 4.6 (1.9, 5.6) | 0.4 (0.0, 0.9) | 0.0 (0.0, 0.0) |
| South Asia | India #1 | 24.3 (20.0, 26.8) | 14.4 (5.8, 18.0) | 9.7 (2.9, 11.9) | 24.5 (5.6, 34.2) | 4.4 (0.3, 5.8) | 1.7 (0.7, 2.2) | 0.6 (0.3, 0.9) | 1.7 (0.8, 2.3) | 2.2 (0.7, 3.3) | 2.7 (0.5, 3.3) | 1.0 (0.0, 3.8) | 0.1 (0.1, 0.2) |
| | India #2 | 34.0 (28.1, 36.9) | 7.9 (3.5, 10.7) | 16.1 (5.4, 19.5) | 14.5 (4.4, 22.0) | 5.1 (0.5, 7.0) | 1.7 (0.7, 2.2) | 0.5 (0.2, 0.8) | 1.3 (0.4, 2.0) | 6.3 (1.3, 9.0) | 1.4 (0.4, 1.7) | 0.0 (0.0, 0.2) | 0.0 (0.0, 0.0) |
| | India #3 | 13.5 (10.1, 15.1) | 15.9 (5.8, 21.1) | 7.4 (2.1, 9.5) | 15.4 (2.7, 24.2) | 7.7 (1.7, 10.3) | 2.4 (0.8, 3.1) | 3.3 (1.1, 5.2) | 6.8 (2.8, 9.0) | 2.3 (0.1, 3.7) | 0.0 (0.0, 0.1) | 1.1 (0.0, 5.0) | 7.0 (6.1, 7.4) |
| | India #4 | 7.2 (5.3, 8.3) | 10.5 (4.5, 14.8) | 7.0 (2.1, 9.2) | 25.7 (13.4, 35.8) | 12.9 (1.6, 18.6) | 2.7 (0.8, 3.6) | 0.0 (0.0, 0.0) | 15.5 (3.4, 21.6) | 1.2 (0.2, 2.0) | 0.7 (0.1, 1.0) | 1.1 (0.0, 5.1) | 0.0 (0.0, 0.0) |
| | India #5* | --- | --- | --- | --- | --- | --- | --- | --- | --- | --- | --- | --- |
| | Pakistan #1* | --- | --- | --- | --- | --- | --- | --- | --- | --- | --- | --- | --- |
| | Pakistan #2* | --- | --- | --- | --- | --- | --- | --- | --- | --- | --- | --- | --- |
| Southeast Asia and Oceania | Fiji | 14.9 (11.9, 16.4) | 18.4 (14.6, 20.0) | 14.4 (5.4, 16.7) | 8.0 (0.1, 10.6) | 5.3 (3.1, 6.7) | 4.4 (3.1, 5.2) | 0.9 (0.6, 1.4) | 2.2 (1.3, 2.7) | 2.2 (0.4, 3.2) | 0.0 (0.0, 0.0) | 2.5 (0.0, 4.9) | 0.0 (0.0, 0.0) |
| | Indonesia | 26.9 (19.4, 30.0) | 9.1 (7.4, 10.3) | 7.8 (6.4, 8.5) | 4.9 (2.0, 6.4) | 1.4 (0.9, 1.6) | 1.0 (0.7, 1.2) | 0.2 (0.1, 0.3) | 2.7 (2.1, 3.3) | 3.4 (1.8, 4.6) | 0.5 (0.0, 0.9) | 0.0 (0.0, 0.0) | 0.6 (0.5, 0.6) |
| | Lao PDR | 40.5 (32.2, 44.4) | 4.6 (3.5, 5.1) | 12.0 (9.8, 13.1) | 1.5 (0.5, 1.8) | 3.8 (2.6, 4.6) | 1.0 (0.6, 1.2) | 1.2 (0.6, 1.8) | 0.4 (0.2, 0.5) | 2.0 (0.9, 2.9) | 2.4 (0.4, 4.0) | 0.2 (0.0, 0.4) | 0.0 (0.0, 0.0) |
| | Myanmar | 59.9 (53.9, 63.4) | 3.7 (2.9, 5.4) | 5.8 (3.6, 8.2) | 15.4 (0.1, 19.2) | 3.1 (0.0, 4.8) | 2.6 (1.1, 3.9) | 1.0 (0.6, 1.2) | 3.9 (1.0, 5.6) | 0.7 (0.0, 2.3) | 0.3 (0.0, 0.4) | 0.0 (0.0, 2.6) | 1.2 (0.9, 1.4) |
| | Philippines* | --- | --- | --- | --- | --- | --- | --- | --- | --- | --- | --- | --- |
| | Viet Nam | 21.8 (16.2, 24.4) | 1.3 (1.0, 1.4) | 19.4 (15.9, 21.6) | 1.3 (0.4, 1.7) | 3.7 (2.9, 4.4) | 1.0 (0.7, 1.3) | 0.1 (0.1, 0.1) | 0.9 (0.5, 1.2) | 3.0 (1.0, 4.0) | 0.2 (0.0, 0.3) | 0.0 (0.0, 0.0) | 0.0 (0.0, 0.0) |
| East Asia | China #1 | 18.5 (13.9, 20.9) | 0.0 (0.0, 0.0) | 5.6 (4.6, 6.7) | 3.1 (1.6, 4.3) | 1.0 (0.8, 1.1) | 0.0 (0.0, 0.0) | 0.0 (0.0, 0.0) | 1.3 (0.8, 1.7) | 0.4 (0.1, 0.5) | 0.9 (0.0, 1.3) | 0.0 (0.0, 0.1) | 0.0 (0.0, 0.0) |
| | China #2 | 21.9 (16.5, 23.8) | 0.0 (0.0, 0.0) | 8.8 (7.2, 9.7) | 1.2 (0.6, 1.6) | 1.9 (1.4, 2.1) | 0.2 (0.1, 0.2) | 0.0 (0.0, 0.0) | 1.4 (0.8, 1.7) | 1.0 (0.4, 1.5) | 1.6 (0.4, 2.5) | 0.0 (0.0, 0.0) | 0.0 (0.0, 0.0) |
| | China #3 | 43.4 (31.9, 48.1) | 0.0 (0.0, 0.0) | 23.4 (19.1, 25.8) | 2.4 (1.2, 3.5) | 1.1 (0.8, 1.4) | 0.0 (0.0, 0.0) | 0.0 (0.0, 0.0) | 1.9 (1.1, 2.4) | 0.8 (0.2, 1.1) | 1.0 (0.1, 1.4) | 0.3 (0.0, 0.7) | 0.0 (0.0, 0.0) |

Attributable fractions are expressed as a percent. Lao PDR=Lao People's Democratic Republic. tEPEC=typical enteropathogenic *E. coli*. ETEC=enterotoxigenic *E. coli*.

*Site did not participate in GPDS during this two-year time period

**S6 Table. All-cause and pathogen-specific attributable incidence per 1000 child-years with 95% confidence intervals of diarrhea hospitalizations in children aged <5 years both overall and by geographic grouping in countries participating in Global Pediatric Diarrhea Surveillance, by two-year time periods during 2017-2022 (Underlying data for Figure 6).**

| | All-cause | Rotavirus | *Shigella* | Norovirus | Adenovirus 40/41 | Sapovirus | ETEC | *Cryptosporidium* | Astrovirus | *C. jejuni/coli* | *Salmonella* | tEPEC | *V. cholerae* |
|---|---|---|---|---|---|---|---|---|---|---|---|---|---|
| **2017-2018** | | | | | | | | | | | | | |
| Overall | 11.9 (10.0, 14.3) | 4.4 (3.5, 5.6) | 1.3 (1.0, 1.6) | 0.7 (0.6, 0.9) | 0.8 (0.6, 1.1) | 0.5 (0.3, 0.7) | 0.6 (0.4, 0.8) | 0.4 (0.3, 0.6) | 0.4 (0.3, 0.5) | 0.2 (0.1, 0.2) | 0.2 (0.1, 0.3) | 0.1 (0.0, 0.1) | 0.1 (0.1, 0.1) |
| Central America | 1.8 (1.4, 2.4) | 0.4 (0.3, 0.5) | 0.3 (0.2, 0.5) | 0.3 (0.1, 0.4) | 0.2 (0.1, 0.3) | 0.1 (0.0, 0.1) | 0.0 (0.0, 0.1) | 0.0 (0.0, 0.0) | 0.1 (0.0, 0.1) | 0.1 (0.0, 0.1) | 0.0 (0.0, 0.0) | 0.0 (0.0, 0.0) | 0.0 (0.0, 0.0) |
| South America | 2.2 (1.7, 2.9) | 0.3 (0.2, 0.4) | 0.3 (0.2, 0.4) | 0.5 (0.4, 0.7) | 0.1 (0.1, 0.2) | 0.1 (0.1, 0.1) | 0.1 (0.0, 0.1) | 0.0 (0.0, 0.0) | 0.0 (0.0, 0.1) | 0.1 (0.0, 0.1) | 0.0 (0.0, 0.0) | 0.0 (0.0, 0.0) | 0.0 (0.0, 0.0) |
| Eastern Europe | 1.4 (0.9, 2.1) | 0.5 (0.3, 0.7) | 0.0 (0.0, 0.0) | 0.1 (0.1, 0.2) | 0.0 (0.0, 0.0) | 0.0 (0.0, 0.1) | 0.0 (0.0, 0.0) | 0.0 (0.0, 0.0) | 0.0 (0.0, 0.0) | 0.1 (0.0, 0.1) | 0.0 (0.0, 0.0) | 0.0 (0.0, 0.0) | 0.0 (0.0, 0.0) |
| Central and Western Asia | 1.1 (0.8, 1.4) | 0.2 (0.2, 0.3) | 0.1 (0.1, 0.2) | 0.1 (0.1, 0.1) | 0.0 (0.0, 0.1) | 0.1 (0.0, 0.1) | 0.0 (0.0, 0.1) | 0.0 (0.0, 0.1) | 0.0 (0.0, 0.1) | 0.0 (0.0, 0.0) | 0.0 (0.0, 0.0) | 0.0 (0.0, 0.0) | 0.0 (0.0, 0.0) |
| West Africa | 43.1 (31.8, 57.7) | 20.8 (14.6, 29.2) | 4.2 (2.4, 6.4) | 1.7 (0.9, 2.5) | 2.3 (0.9, 3.5) | 1.9 (0.6, 2.9) | 2.5 (1.0, 3.8) | 1.5 (0.7, 2.5) | 1.5 (0.7, 2.4) | 0.2 (0.1, 0.2) | 0.9 (0.2, 1.5) | 0.2 (0.1, 0.3) | 0.0 (0.0, 0.1) |
| East and Southern Africa | 26.8 (21.6, 33.2) | 5.5 (4.5, 6.7) | 3.9 (2.8, 5.1) | 1.9 (1.3, 2.5) | 1.3 (0.8, 1.8) | 1.2 (0.9, 1.6) | 1.7 (1.2, 2.3) | 1.6 (1.1, 2.1) | 0.7 (0.5, 1.1) | 0.6 (0.2, 0.9) | 0.2 (0.1, 0.2) | 0.4 (0.1, 0.8) | 0.2 (0.2, 0.2) |
| South Asia | 4.9 (3.4, 7.1) | 1.2 (0.8, 1.7) | 0.6 (0.3, 1.0) | 0.5 (0.2, 0.8) | 0.8 (0.4, 1.4) | 0.3 (0.1, 0.4) | 0.1 (0.0, 0.1) | 0.1 (0.0, 0.1) | 0.2 (0.1, 0.3) | 0.2 (0.1, 0.3) | 0.1 (0.0, 0.1) | 0.1 (0.0, 0.1) | 0.1 (0.1, 0.2) |
| Southeast Asia and Oceania | 10.3 (8.2, 13.1) | 3.3 (2.6, 4.2) | 0.6 (0.4, 0.9) | 1.0 (0.8, 1.3) | 0.6 (0.3, 0.8) | 0.2 (0.1, 0.3) | 0.1 (0.1, 0.2) | 0.0 (0.0, 0.0) | 0.3 (0.2, 0.4) | 0.3 (0.2, 0.4) | 0.0 (0.0, 0.1) | 0.0 (0.0, 0.1) | 0.1 (0.0, 0.1) |
| East Asia | 0.7 (0.4, 1.0) | 0.2 (0.1, 0.3) | 0.0 (0.0, 0.0) | 0.1 (0.1, 0.1) | 0.0 (0.0, 0.0) | 0.0 (0.0, 0.0) | 0.0 (0.0, 0.0) | 0.0 (0.0, 0.0) | 0.0 (0.0, 0.0) | 0.0 (0.0, 0.0) | 0.0 (0.0, 0.0) | 0.0 (0.0, 0.0) | 0.0 (0.0, 0.0) |
| **2019-2020** | | | | | | | | | | | | | |
| Overall | 9.9 (8.3, 11.9) | 3.2 (2.3, 4.3) | 0.9 (0.7, 1.1) | 0.7 (0.5, 0.8) | 1.2 (0.6, 1.8) | 0.4 (0.2, 0.5) | 0.3 (0.3, 0.4) | 0.2 (0.1, 0.3) | 0.2 (0.1, 0.2) | 0.1 (0.1, 0.2) | 0.1 (0.0, 0.1) | 0.1 (0.0, 0.2) | 0.1 (0.1, 0.1) |
| Central America | 1.7 (1.2, 2.3) | 0.4 (0.3, 0.5) | 0.2 (0.1, 0.3) | 0.4 (0.2, 0.5) | 0.1 (0.1, 0.2) | 0.1 (0.0, 0.1) | 0.0 (0.0, 0.0) | 0.0 (0.0, 0.0) | 0.1 (0.0, 0.1) | 0.0 (0.0, 0.0) | 0.0 (0.0, 0.0) | 0.0 (0.0, 0.0) | 0.0 (0.0, 0.0) |
| South America | 1.9 (1.4, 2.4) | 0.2 (0.2, 0.3) | 0.3 (0.2, 0.4) | 0.4 (0.3, 0.5) | 0.1 (0.1, 0.2) | 0.1 (0.1, 0.1) | 0.1 (0.0, 0.1) | 0.0 (0.0, 0.0) | 0.0 (0.0, 0.0) | 0.0 (0.0, 0.1) | 0.0 (0.0, 0.0) | 0.0 (0.0, 0.0) | 0.0 (0.0, 0.0) |
| Eastern Europe | 1.3 (0.8, 1.9) | 0.3 (0.2, 0.5) | 0.0 (0.0, 0.0) | 0.2 (0.1, 0.2) | 0.1 (0.0, 0.2) | 0.0 (0.0, 0.0) | 0.0 (0.0, 0.0) | 0.0 (0.0, 0.0) | 0.0 (0.0, 0.0) | 0.1 (0.0, 0.1) | 0.0 (0.0, 0.0) | 0.0 (0.0, 0.0) | 0.0 (0.0, 0.0) |
| Central and Western Asia | 2.1 (1.5, 2.9) | 0.5 (0.3, 0.7) | 0.3 (0.2, 0.4) | 0.2 (0.2, 0.4) | 0.1 (0.1, 0.2) | 0.1 (0.1, 0.2) | 0.0 (0.0, 0.0) | 0.0 (0.0, 0.1) | 0.1 (0.0, 0.1) | 0.0 (0.0, 0.0) | 0.0 (0.0, 0.0) | 0.0 (0.0, 0.1) | 0.0 (0.0, 0.0) |
| West Africa | 35.4 (26.1, 47.2) | 16.2 (10.1, 23.1) | 2.0 (1.0, 2.9) | 2.2 (1.0, 3.2) | 5.0 (1.1, 8.5) | 1.3 (0.3, 2.1) | 0.8 (0.4, 1.2) | 0.6 (0.3, 0.9) | 0.4 (0.2, 0.6) | 0.3 (0.2, 0.4) | 0.2 (0.1, 0.4) | 0.3 (0.1, 0.7) | 0.0 (0.0, 0.0) |
| East and Southern Africa | 20.0 (16.0, 24.9) | 3.5 (2.9, 4.2) | 3.1 (2.2, 4.1) | 1.5 (0.9, 2.0) | 1.4 (0.7, 1.9) | 0.9 (0.6, 1.2) | 1.0 (0.7, 1.4) | 0.7 (0.5, 1.0) | 0.6 (0.4, 0.9) | 0.4 (0.2, 0.7) | 0.0 (0.0, 0.0) | 0.2 (0.1, 0.5) | 0.0 (0.0, 0.0) |
| South Asia | 4.8 (3.6, 6.3) | 0.6 (0.5, 0.8) | 0.6 (0.3, 0.9) | 0.3 (0.1, 0.5) | 0.6 (0.4, 0.9) | 0.2 (0.1, 0.3) | 0.2 (0.1, 0.4) | 0.1 (0.1, 0.1) | 0.1 (0.1, 0.1) | 0.1 (0.0, 0.2) | 0.0 (0.0, 0.1) | 0.1 (0.0, 0.2) | 0.2 (0.1, 0.2) |
| Southeast Asia and Oceania | 11.3 (8.7, 14.5) | 3.7 (2.7, 5.0) | 0.3 (0.2, 0.5) | 0.9 (0.6, 1.3) | 1.1 (0.0, 1.8) | 0.2 (0.1, 0.3) | 0.1 (0.0, 0.2) | 0.0 (0.0, 0.0) | 0.1 (0.0, 0.1) | 0.2 (0.0, 0.4) | 0.1 (0.0, 0.2) | 0.0 (0.0, 0.2) | 0.0 (0.0, 0.0) |
| East Asia | 0.6 (0.4, 0.9) | 0.1 (0.1, 0.2) | 0.0 (0.0, 0.0) | 0.1 (0.1, 0.1) | 0.0 (0.0, 0.0) | 0.0 (0.0, 0.0) | 0.0 (0.0, 0.0) | 0.0 (0.0, 0.0) | 0.0 (0.0, 0.0) | 0.0 (0.0, 0.0) | 0.0 (0.0, 0.0) | 0.0 (0.0, 0.0) | 0.0 (0.0, 0.0) |
| **2021-2022** | | | | | | | | | | | | | |
| Overall | 8.8 (7.4, 10.5) | 2.3 (1.8, 3.0) | 0.9 (0.7, 1.1) | 0.6 (0.4, 0.8) | 0.6 (0.4, 0.8) | 0.2 (0.1, 0.3) | 0.2 (0.2, 0.3) | 0.2 (0.1, 0.2) | 0.1 (0.1, 0.1) | 0.1 (0.1, 0.2) | 0.1 (0.1, 0.1) | 0.1 (0.0, 0.1) | 0.1 (0.1, 0.2) |
| Central America | 1.7 (1.3, 2.3) | 0.2 (0.2, 0.3) | 0.2 (0.1, 0.2) | 0.3 (0.1, 0.4) | 0.1 (0.0, 0.2) | 0.1 (0.0, 0.1) | 0.0 (0.0, 0.1) | 0.0 (0.0, 0.0) | 0.0 (0.0, 0.1) | 0.0 (0.0, 0.1) | 0.0 (0.0, 0.0) | 0.0 (0.0, 0.0) | 0.0 (0.0, 0.0) |
| South America | 1.5 (1.1, 1.9) | 0.1 (0.1, 0.2) | 0.2 (0.1, 0.2) | 0.3 (0.2, 0.4) | 0.1 (0.1, 0.1) | 0.0 (0.0, 0.1) | 0.0 (0.0, 0.1) | 0.0 (0.0, 0.0) | 0.0 (0.0, 0.0) | 0.1 (0.0, 0.1) | 0.0 (0.0, 0.0) | 0.0 (0.0, 0.0) | 0.0 (0.0, 0.0) |
| Eastern Europe | 2.0 (1.4, 3.0) | 0.5 (0.3, 0.7) | 0.0 (0.0, 0.0) | 0.3 (0.2, 0.4) | 0.1 (0.1, 0.2) | 0.0 (0.0, 0.0) | 0.0 (0.0, 0.0) | 0.0 (0.0, 0.0) | 0.0 (0.0, 0.0) | 0.1 (0.0, 0.1) | 0.0 (0.0, 0.0) | 0.0 (0.0, 0.0) | 0.0 (0.0, 0.0) |
| Central and Western Asia | 2.0 (1.4, 2.8) | 0.3 (0.2, 0.5) | 0.2 (0.1, 0.3) | 0.2 (0.2, 0.4) | 0.1 (0.0, 0.1) | 0.1 (0.1, 0.1) | 0.0 (0.0, 0.0) | 0.0 (0.0, 0.1) | 0.0 (0.0, 0.0) | 0.0 (0.0, 0.0) | 0.0 (0.0, 0.0) | 0.0 (0.0, 0.0) | 0.0 (0.0, 0.0) |
| West Africa | 29.1 (21.0, 39.6) | 10.5 (7.0, 15.0) | 1.4 (0.7, 2.0) | 2.1 (0.9, 3.1) | 2.7 (1.1, 4.3) | 0.6 (0.2, 1.0) | 0.4 (0.2, 0.6) | 0.6 (0.2, 1.0) | 0.3 (0.2, 0.5) | 0.1 (0.0, 0.1) | 0.1 (0.0, 0.1) | 0.2 (0.0, 0.4) | 0.0 (0.0, 0.0) |
| East and Southern Africa | 18.0 (14.5, 22.5) | 3.6 (3.0, 4.4) | 3.3 (2.4, 4.5) | 1.0 (0.7, 1.4) | 0.9 (0.5, 1.3) | 0.6 (0.4, 0.8) | 1.0 (0.7, 1.2) | 0.7 (0.5, 1.0) | 0.3 (0.2, 0.4) | 0.5 (0.2, 0.7) | 0.2 (0.0, 0.4) | 0.2 (0.1, 0.4) | 0.0 (0.0, 0.0) |
| South Asia | 4.3 (3.2, 5.8) | 0.7 (0.5, 0.9) | 0.7 (0.3, 1.1) | 0.2 (0.1, 0.4) | 0.2 (0.1, 0.3) | 0.1 (0.0, 0.2) | 0.1 (0.1, 0.2) | 0.0 (0.0, 0.1) | 0.0 (0.0, 0.1) | 0.1 (0.0, 0.2) | 0.1 (0.0, 0.1) | 0.0 (0.0, 0.1) | 0.2 (0.1, 0.2) |
| Southeast Asia and Oceania | 10.5 (7.9, 13.9) | 2.2 (1.5, 3.0) | 0.6 (0.3, 0.9) | 1.7 (1.1, 2.4) | 0.3 (0.1, 0.5) | 0.3 (0.2, 0.5) | 0.1 (0.1, 0.2) | 0.0 (0.0, 0.0) | 0.2 (0.1, 0.3) | 0.5 (0.2, 0.8) | 0.2 (0.1, 0.4) | 0.0 (0.0, 0.1) | 0.8 (0.5, 1.2) |
| East Asia | 0.5 (0.3, 0.8) | 0.1 (0.1, 0.1) | 0.0 (0.0, 0.0) | 0.1 (0.0, 0.1) | 0.0 (0.0, 0.1) | 0.0 (0.0, 0.0) | 0.0 (0.0, 0.0) | 0.0 (0.0, 0.0) | 0.0 (0.0, 0.0) | 0.0 (0.0, 0.0) | 0.0 (0.0, 0.0) | 0.0 (0.0, 0.0) | 0.0 (0.0, 0.0) |

tEPEC=typical enteropathogenic *E. coli*. ETEC=heat-stable enterotoxigenic *E. coli*

**S1 Fig. Prevalence of pathogens including in the attribution modelling at cycle threshold cutoffs of 35 and 30 in children <5 years hospitalized with diarrhea and enrolled in Global Pediatric Diarrhea Surveillance, by site and two-year time period from 2017-2022.**

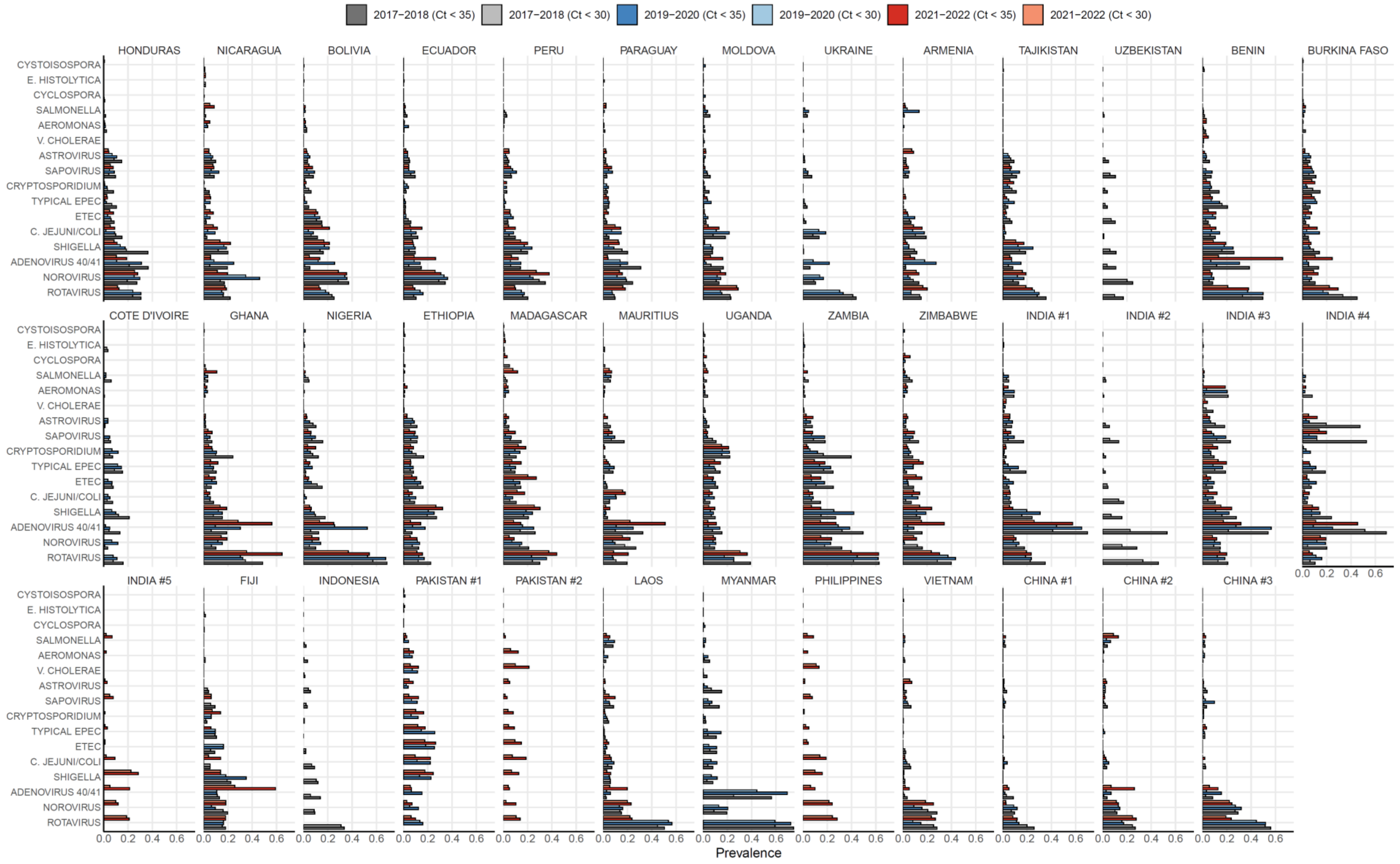
